# Artificial Intelligence-based Automated Echocardiographic Analysis and the Workflow of Sonographers: A randomized crossover trial

**DOI:** 10.1101/2025.08.20.25334115

**Authors:** Akira Sakamoto, Nobuyuki Kagiyama, Eiichiro Sato, Yutaka Nakamura, Azusa Murata, Tomohiro Kaneko, Sakiko Miyazaki, Yuko Ashikawa, Kenichi Sugihara, Tohru Minamino

## Abstract

**Background:** This randomized crossover trial aimed to evaluate whether an artificial intelligence (AI)-based automatic analysis tool for echocardiography could improve the daily workflow of sonographers in real-world clinical practice.

**Methods:** A single-center randomized crossover trial was conducted with four certified sonographers. Each study day, the use of AI-based automatic echocardiography analysis was randomly assigned: either AI assistance (AI days) or manual workflow (non-AI days). The AI tool automatically analyzed echocardiographic images and provided measurements, enabling sonographers to focus on verifying AI-generated values. Expert echocardiologists reviewed and finalized all reports. The primary endpoint was examination efficiency, measured by examination time per patient and the number of examinations performed per day. Secondary endpoints included sonographer fatigue, the number of analyzed echocardiographic parameters, and image quality.

**Results:** A total of 585 patients were scanned over 38 study days (AI days: 317; non-AI days: 268) between Jan 30 and Mar 26, 2024. Baseline characteristics were comparable between groups. AI assistance significantly reduced examination time (13.0 ± 3.5 minutes vs. 14.3 ± 4.2 minutes, p<0.001) and increased the average number of daily examinations (16.7 ± 2.5 vs. 14.1 ± 2.5, p=0.003). Despite the higher workload, sonographers reported lower mental fatigue scores on AI days (4.1 ± 1.1 vs. 4.7 ± 0.6, p=0.039). The number of echocardiographic parameters analyzed per examination increased 3.4-fold on AI days (85 ± 12 vs. 25 ± 1, p<0.001). Differences between AI-generated measurements and final expert-endorsed values were within acceptable clinical limits for 90% of parameters. Notably, image quality significantly improved on AI days (p<0.001).

**Conclusions:** This real-world randomized trial demonstrated that AI-based echocardiographic analysis can enhance workflow efficiency, reduce sonographer fatigue, and improve image quality without compromising diagnostic integrity. Integrating AI into clinical practice holds promise for optimizing high-volume echocardiography workflows.

**Highlights:** Artificial intelligence (AI)-assisted echocardiography improved sonographer workflow efficiency.

AI assistance significantly reduced examination time and increased daily scan volume. Sonographer mental fatigue was lower despite the increased workload on AI days.

AI integration markedly increased the number of analyzed echocardiographic parameters.

AI-enhanced workflow improved echocardiographic image quality without compromising accuracy.

**Graphical abstract:** This first-ever trial to randomize the use of an AI-based automated tool on a daily basis revealed that AI significantly enhanced the efficiency of screening echocardiography, reducing examination time despite a 3.4-fold increase in the number of parameters measured. This improved efficiency increased the number of examinations per day without increasing sonographers’ fatigue; in fact, it mitigated mental fatigue. Furthermore, being freed from the need to perform time-consuming measurements allowed sonographers to focus on image acquisition, which led to improved image quality.

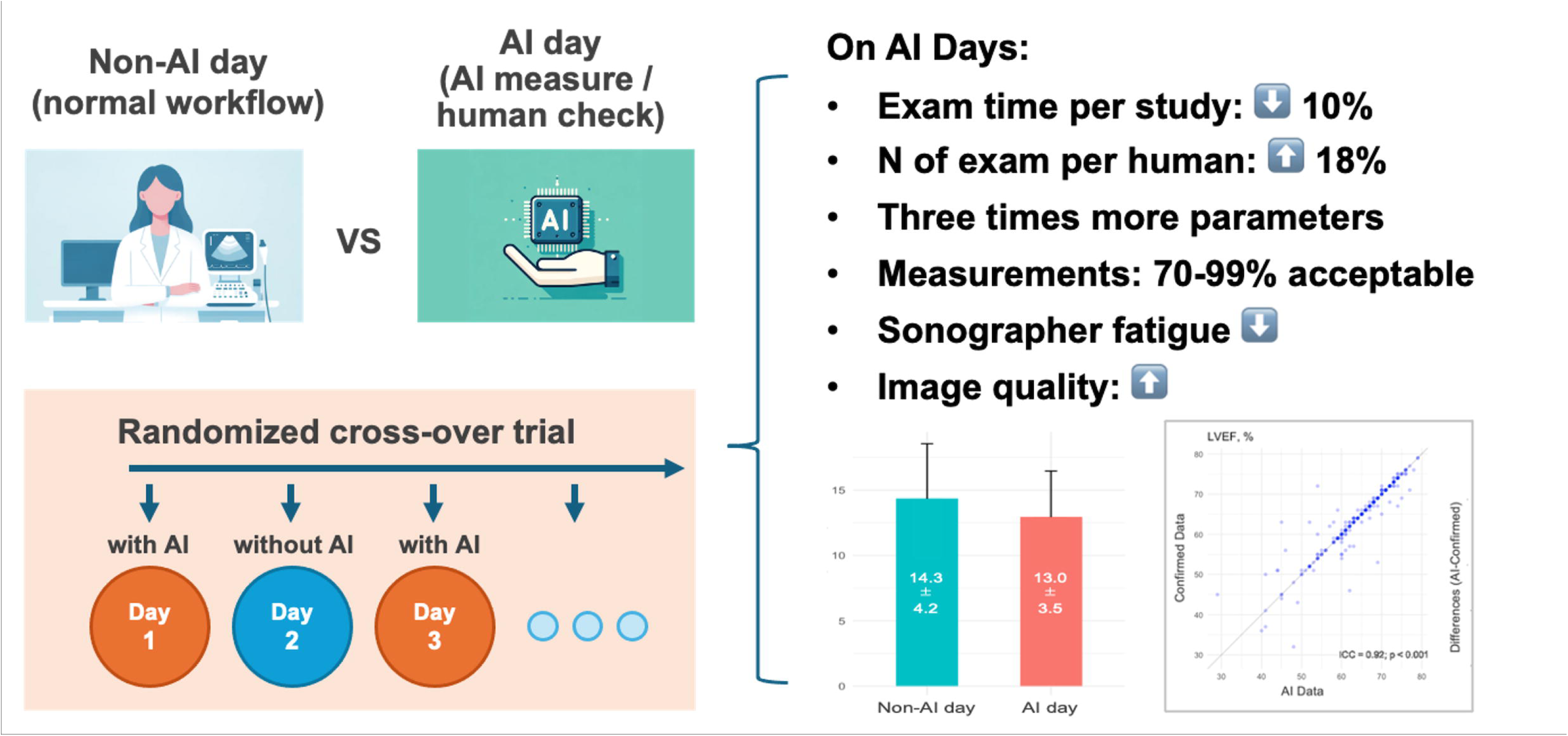

## Introduction

Echocardiography is a cornerstone, non-invasive diagnostic tool in cardiovascular medicine, widely used for its ability to provide detailed insights into cardiac structure and function. Over recent years, the increasing prevalence of cardiovascular disease has driven a corresponding rise in the demand for echocardiographic examinations ^1^. This trend has been particularly pronounced in the context of screening echocardiography, which often involves repetitive, standardized tasks. Such a large number of examinations in routine workflows can diminish sonographers’ motivation, contributing to fatigue and elevated rates of burnout ^2^. Addressing this issue, optimizing the efficiency of echocardiographic workflows has become an urgent priority, especially in countries like Japan, where the number of echocardiographic examinations per capita is 3.5 times higher than in the United States ^3^.

In recent years, artificial intelligence (AI) has advanced rapidly, with significant potential applications in healthcare ^4–6^. In echocardiography, AI excels in image recognition and has shown great promise in the automatic classification and analysis of echocardiographic images ^7, 8^. Recent studies have validated AI’s capability to automate the analysis of most routine echocardiographic parameters, reducing analysis time while maintaining high reproducibility ^9–12^. These tools offer the potential to streamline sonographers’ workflows by reducing time spent on routine measurements, thereby allowing greater focus on more complex tasks, such as managing challenging patient cases or interpreting echocardiographic findings ^13^. However, much of the existing research has been conducted retrospectively or in controlled experimental environments, leaving questions regarding AI’s impact on workflow efficiency in real-world clinical settings.

To address this gap, we conducted a randomized crossover trial to evaluate whether an AI-based automatic analysis tool for echocardiography could streamline the daily workflow of sonographers in a real-world clinical environment.

## Methods

### Study design

This single-center, single-blinded, randomized crossover study was conducted at Juntendo University Hospital (Tokyo, Japan) between January 2024 and March 2024. Certified clinical sonographers, credentialed by the Japan Society of Ultrasonics in Medicine, were enrolled in the study after providing written informed consent. The study protocol received approval from the Research Ethics Committee of Faculty of Medicine, Juntendo University (approval number: E23-0161-M01), and the study was registered in the University Hospital Medical Information Network Clinical Trials Registry (identification number: UMIN000053259) prior to participant enrollment. All procedures adhered to the principles outlined in the Declaration of Helsinki, and the CONSORT checklist ^14^ is available in the Supplemental Table 1.

### Randomization of AI and Non-AI Days

The study employed a randomized crossover design, wherein echocardiographic examinations were conducted either with AI assistance (AI days) or without AI assistance (non-AI days). The echocardiography laboratory comprised eight booths, each staffed by a sonographer on a daily basis. One specific booth with a dedicated ultrasound machine (Epic Elite; Philips Healthcare, Netherlands) was designated for the randomized AI or non-AI workflow throughout the study, while the remaining booths followed standard procedures. The allocation of AI and non-AI days for this designated booth was determined using computer-generated block randomization, considering the day of the week. A custom software program was used to manage the randomization, and sonographers accessed the program each morning to determine whether that day’s examinations would involve AI assistance.

The laboratory handled two types of examinations: “detailed” echocardiography for patients with known cardiac diseases, and “screening” echocardiography for preoperative assessments, chest pain evaluations, and for patients without a known history of cardiovascular disease. However, the study booth focused exclusively on screening examinations, as these repetitive and formulaic tasks are precisely the type of examinations that AI is expected to streamline.

## 1. AI-Assisted workflow (AI days)

On AI days, sonographers utilized an automated AI analysis tool, specifically the FDA-approved US2.ai platform (Us2.ai, Singapore). This platform automatically measured key echocardiographic parameters (detailed in the Supplemental Table 2), eliminating the need for manual measurements in most cases ^11^. Images were uploaded to an on-premise server, and AI analysis results were sent back to the reporting system (ISCV; Philips Healthcare, Netherlands) within approximately 2 minutes. Sonographers then reviewed these AI-generated results, which included trace lines overlaid on the images to illustrate the AI’s analysis (Supplemental Figure 1). Certain parameters, such as the diameters of the left atrium, aortic root, and inferior vena cava, were not measured by the AI and required manual measurement. If any AI-generated measurements or tracings were considered inaccurate, the sonographer remeasured the parameters manually on the workstation. Both AI and non-AI reports were reviewed and finalized by expert echocardiologists, who were blinded to the AI status, ensuring all reports were suitable for clinical use.

## 2. Standard workflow (Non-AI days)

On non-AI days, sonographers performed echocardiographic examinations without AI assistance, following their routine clinical procedures. Sonographers were allowed to select their preferred method for conducting measurements, whether directly on the echocardiography machine or at the workstation post-scan. All measurements adhered to established guidelines ^15, 16^.

### Study endpoints

The primary endpoints of this study were the efficiency of echocardiographic examinations, measured by the total time required for scanning, analysis, and reporting for each patient, as well as the total number of patients scanned and reported on by each sonographer during a typical workday (from 9 a.m. to 5 p.m.).

Secondary endpoints included (1) sonographers’ mental and physical fatigue at the end of each workday, measured using a daily questionnaire shown in Supplemental Table 3, (2) the number of echocardiographic parameters analyzed per examination, (3) the quality of echocardiographic images, and (4) AI’s performance, assessed by the rate at which the AI analyzed acquired images and the concordance between AI’s initial measurements and the final values endorsed by expert echocardiologists.

The image quality was evaluated by two blinded echocardiologists who assessed five standard views: parasternal long-axis, short-axis, apical 4-chamber, 3-chamber, and 2-chamber views. Each view was graded on a 3-point scale (Poor, > 5 segments poorly visible; Good, 3–5 segments poorly visible; and Excellent, only 0–2 segments poorly visible throughout the cardiac cycle), adapted from a previous report ^9, 17^.

### Sample size calculation

The required number of study days was determined based on a sample size calculation. Previous research has demonstrated that using this AI system results in a 70% reduction in analysis time ^9^. However, considering the complexity of real-world processes in an echocardiographic laboratory, we conservatively estimated an overall efficiency improvement of 20%, corresponding to an increase in the number of examinations per day from approximately 10 ± 2 to 12 ± 2. The sample size calculation indicated that 16.7 days per arm would be necessary to detect this difference (Alpha = 0.05, Beta = 0.8). To account for an estimated 10% data attrition, we ultimately set the study to include 19 days of examinations per arm.

### Statistical analysis

Data are presented as mean ± standard deviation or medians [1st and 3rd interquartile ranges] for continuous variables, as appropriate, and as frequencies (%) for categorical variables. Group differences were evaluated using Welch’s t-test or Mann-Whitney U test for continuous variables and the chi-square test or Fisher’s exact test for categorical variables. Bland-Altman plots were created to visualize systematic errors between the AI’s initial measurements and the final report values, with limits of agreement defined by the mean bias ± 1.96 standard deviations. Inter-rater reproducibility of the image quality assessments between the two echocardiologists was evaluated using intraclass correlation coefficients (ICC (2,1)). All statistical analyses were performed using R (version 4.4.1; The R Foundation for Statistical Computing, Vienna, Austria). For all analyses, a two-tailed p-value <0.05 was considered statistically significant.

## Results

### Sonographers and examination characteristics

During the study period, four sonographers (average experience in echocardiography 9.0 ± 4.4 years, all female) participated in the study, scanning a total of 585 patients over 38 days, with 19 days each allocated to AI days and non-AI days. These sonographers had experienced the AI-based workflow for one month before starting the study. As summarized in Table 1, the characteristics recorded in the sonographers’ reports, such as patient gender, age, body size, and echocardiographic parameters, were well balanced between AI and non-AI days. Most scans were performed on patients in sinus rhythm, with less than 4% showing atrial fibrillation. The echocardiographic parameters remained largely within normal ranges ^18, 19^ and the prevalence of valvular heart disease was minimal, reflecting that the examinations were for screening purpose.

**Table 1.**
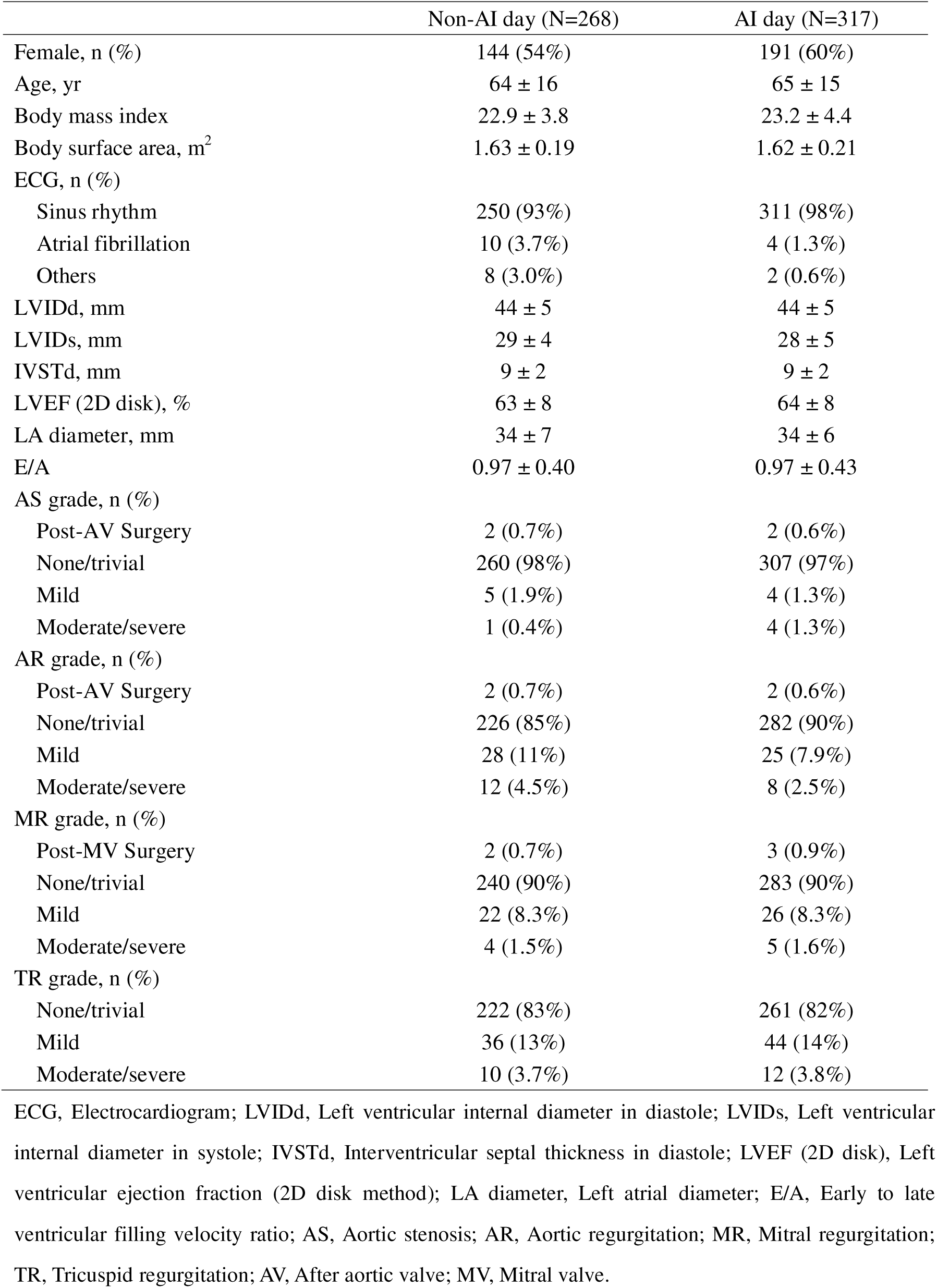
Scan Characteristics on AI and Non-AI Days

### Examination efficiency and sonographer workload

The primary endpoint of average examination time was reduced to 13.0 ± 3.5 minutes on AI days, compared to 14.3 ± 4.2 minutes on non-AI days (p < 0.001) (Figure 1A). As a result, the number of echocardiography performed per sonographer increased significantly on AI days, averaging 16.7 ± 2.5 examinations per day, compared to 14.1 ± 2.5 on non-AI days (p = 0.003) (Figure 1B). Over the course of the study, 317 examinations were performed on AI days and 268 on non-AI days. Additionally, the number of echocardiographic parameters analyzed per examination increased 3.4-fold on AI days compared to non-AI days (85 ± 12 vs. 25 ± 1, p < 0.001; Figure 2 A). Notably, left ventricular (LV) and atrial strain were analyzed in 90% and 80% of AI days, respectively, whereas these parameters were never analyzed on non-AI days.

**Figure 1.**
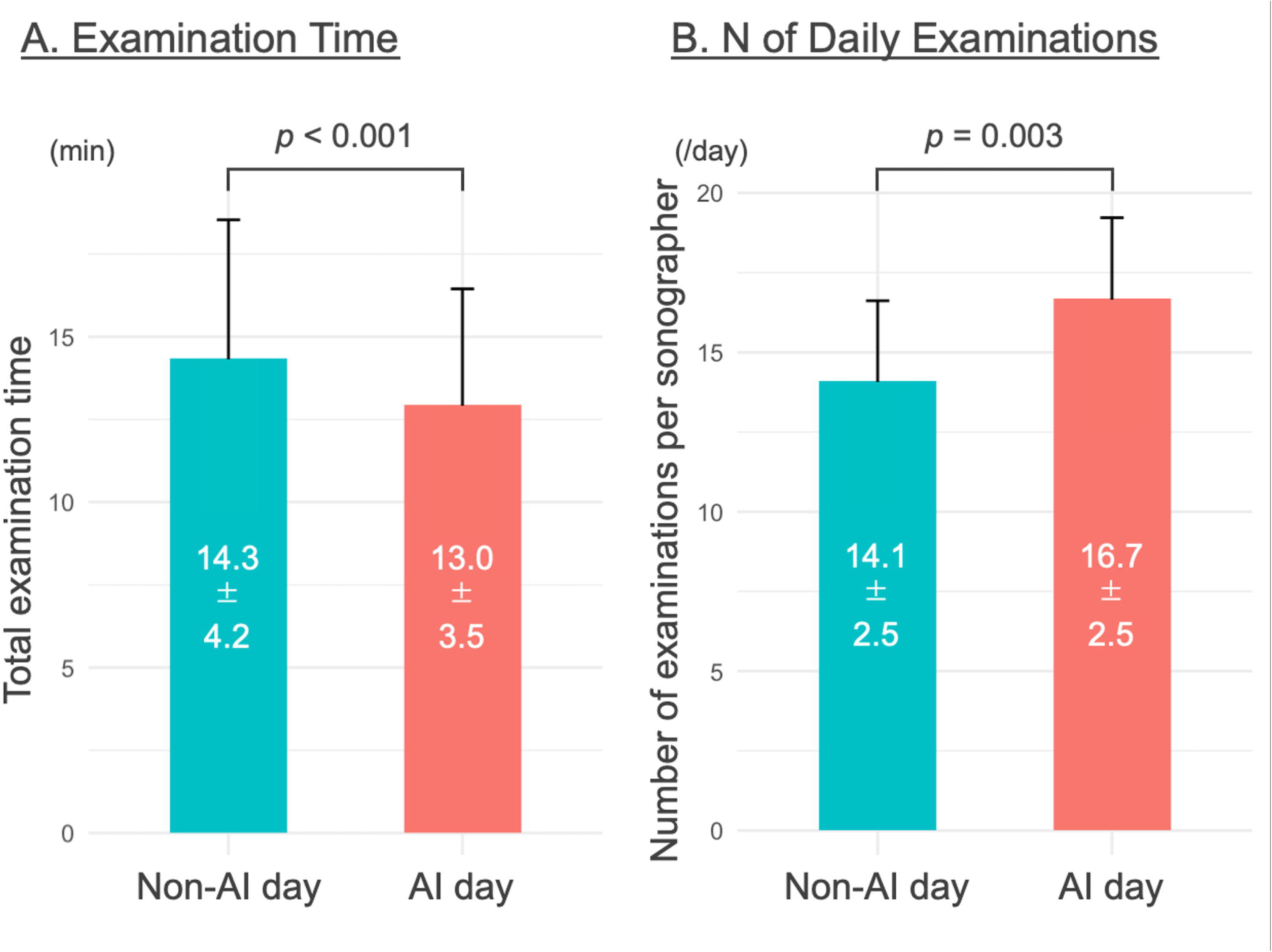
Primary Endpoints of Examination Efficiency On AI days, the average examination time was reduced compared to non-AI days. As a result, the number of echocardiograms performed per sonographer increased significantly on AI days.

**Figure 2.**
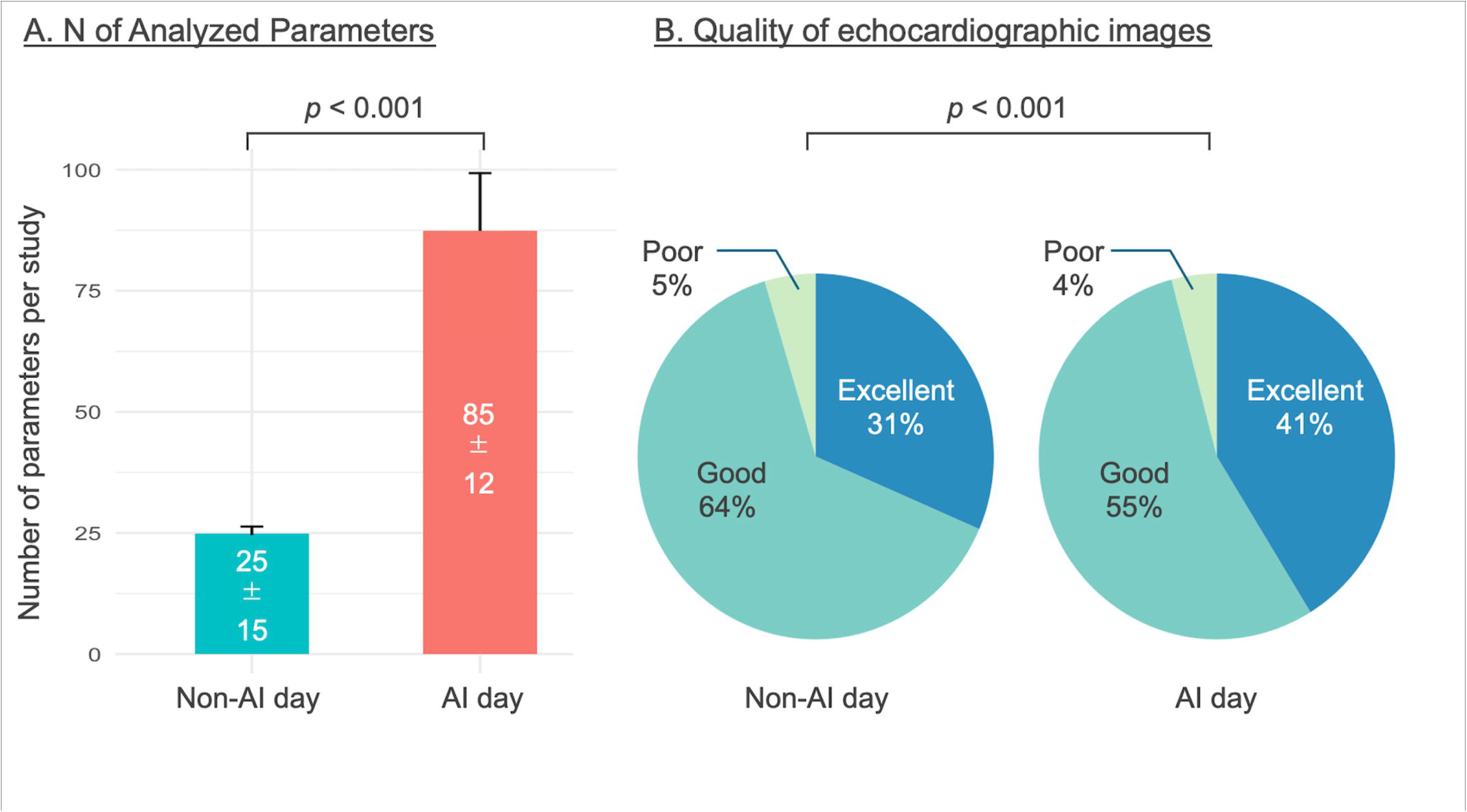
Number of Parameters and Image Quality On AI days, the number of analyzed parameters increased by up to 3.4-fold compared to non-AI days. Additionally, image quality was significantly higher on AI days.

Despite the increase in the number of examinations, the Likert scale-based questionnaire results indicated that mental fatigue experienced by sonographers was significantly lower on AI days compared to non-AI days (4.1 ± 1.1 vs. 4.7 ± 0.6, p = 0.039). The questionnaire also revealed that sonographers’ mental fatigue (4.0 ± 0.9 vs. 4.5 ± 0.8, p = 0.088) and perception of task complexity was numerically lower on AI days (3.7 ± 1.0 vs. 4.2 ± 0.8, p = 0.21), and that their perceived work speed was numerically faster on AI days (3.9 ± 1.1 vs. 3.3 ± 1.6, p = 0.26).

### Improved image quality on AI days

We compared the quality of images acquired on AI days with those obtained on non-AI days to test whether, on AI-assisted days, sonographers could dedicate more attention to image acquisition, resulting in higher-quality echocardiographic images. Two experienced echocardiologists, who were not involved in the report finalization and were blinded to the examination allocation and study results, independently reviewed all images using the evaluation methods outlined in the Methods section. Inter-observer variability tested on 150 images showed excellent agreement, with an ICC of 0.82. A total of 2,925 images were evaluated and categorized as poor, good, or excellent quality. The results, shown in Figure 2B, demonstrated that percentage of images rated as excellent was significantly higher on AI days, with 41% of images achieving this rating compared to 31% on non-AI days (p < 0.001).

### AI’s performance in real-world clinical practice

To assess the AI’s performance in this trial, the rate at which the AI analyzed acquired images and the concordance between AI’s initial measurements and the final values endorsed by expert echocardiologists were analyzed, as summarized in Table 2. Overall, the AI successfully analyzed most parameters in approximately 95% of the cases from available images, while transmitral inflow parameters and left atrial volume index showed an analysis rate of 80–85%.

**Table 2.**
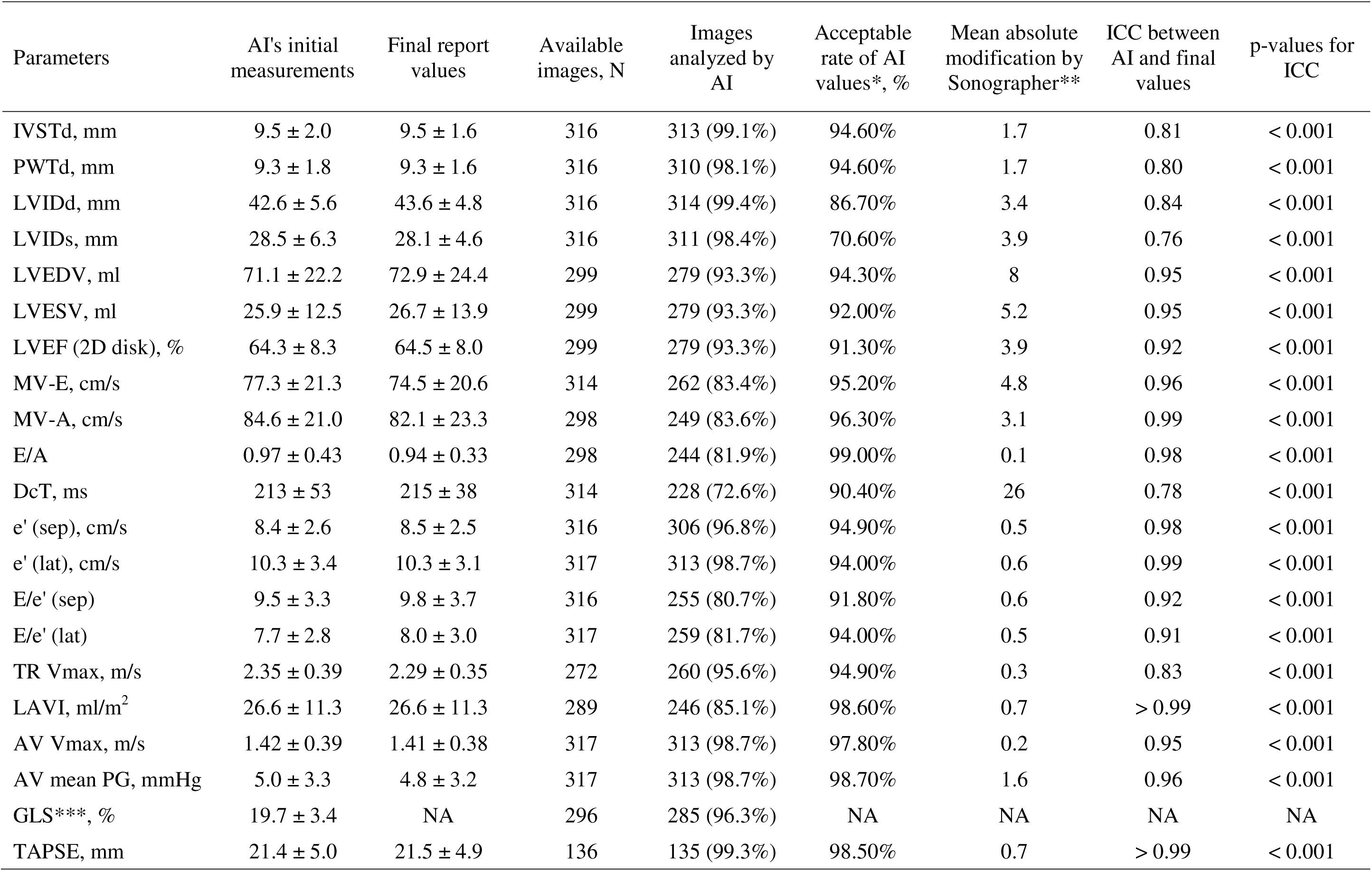
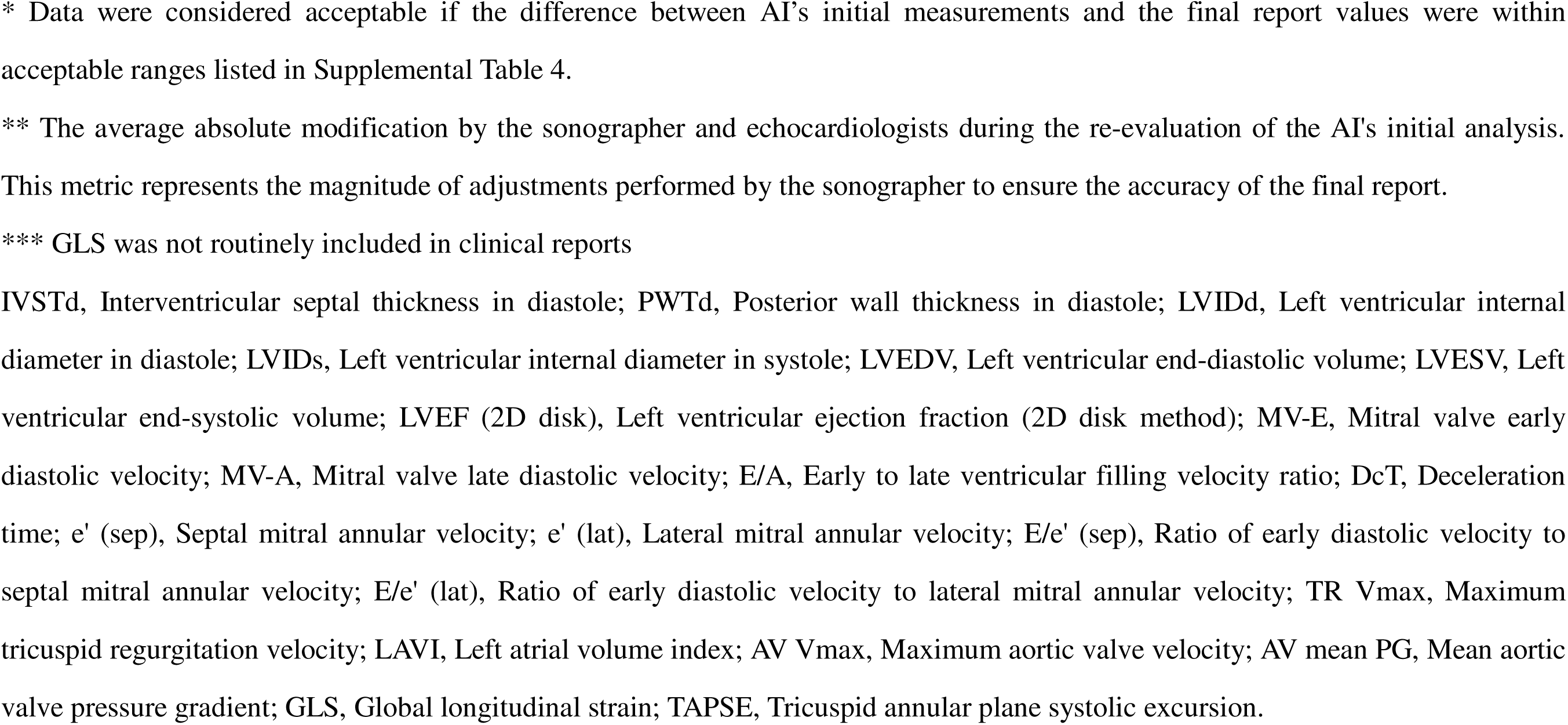
Concordance between AI’s Initial Measurements and Final Report Values

| Parameters | AI's initial measurements | Final report values | Available images, N | Images analyzed by AI | Acceptable rate of AI values*, % | Mean absolute modification by Sonographer** | ICC between AI and final values | p-values for ICC |
| --- | --- | --- | --- | --- | --- | --- | --- | --- |
| IVSTd, mm | 9.5 ± 2.0 | 9.5 ± 1.6 | 316 | 313 (99.1%) | 94.60% | 1.7 | 0.81 | < 0.001 |
| PWTd, mm | 9.3 ± 1.8 | 9.3 ± 1.6 | 316 | 310 (98.1%) | 94.60% | 1.7 | 0.80 | < 0.001 |
| LVIDd, mm | 42.6 ± 5.6 | 43.6 ± 4.8 | 316 | 314 (99.4%) | 86.70% | 3.4 | 0.84 | < 0.001 |
| LVIDs, mm | 28.5 ± 6.3 | 28.1 ± 4.6 | 316 | 311 (98.4%) | 70.60% | 3.9 | 0.76 | < 0.001 |
| LVEDV, ml | 71.1 ± 22.2 | 72.9 ± 24.4 | 299 | 279 (93.3%) | 94.30% | 8 | 0.95 | < 0.001 |
| LVESV, ml | 25.9 ± 12.5 | 26.7 ± 13.9 | 299 | 279 (93.3%) | 92.00% | 5.2 | 0.95 | < 0.001 |
| LVEF (2D disk), % | 64.3 ± 8.3 | 64.5 ± 8.0 | 299 | 279 (93.3%) | 91.30% | 3.9 | 0.92 | < 0.001 |
| MV-E, cm/s | 77.3 ± 21.3 | 74.5 ± 20.6 | 314 | 262 (83.4%) | 95.20% | 4.8 | 0.96 | < 0.001 |
| MV-A, cm/s | 84.6 ± 21.0 | 82.1 ± 23.3 | 298 | 249 (83.6%) | 96.30% | 3.1 | 0.99 | < 0.001 |
| E/A | 0.97 ± 0.43 | 0.94 ± 0.33 | 298 | 244 (81.9%) | 99.00% | 0.1 | 0.98 | < 0.001 |
| DcT, ms | 213 ± 53 | 215 ± 38 | 314 | 228 (72.6%) | 90.40% | 26 | 0.78 | < 0.001 |
| e' (sep), cm/s | 8.4 ± 2.6 | 8.5 ± 2.5 | 316 | 306 (96.8%) | 94.90% | 0.5 | 0.98 | < 0.001 |
| e' (lat), cm/s | 10.3 ± 3.4 | 10.3 ± 3.1 | 317 | 313 (98.7%) | 94.00% | 0.6 | 0.99 | < 0.001 |
| E/e' (sep) | 9.5 ± 3.3 | 9.8 ± 3.7 | 316 | 255 (80.7%) | 91.80% | 0.6 | 0.92 | < 0.001 |
| E/e' (lat) | 7.7 ± 2.8 | 8.0 ± 3.0 | 317 | 259 (81.7%) | 94.00% | 0.5 | 0.91 | < 0.001 |
| TR Vmax, m/s | 2.35 ± 0.39 | 2.29 ± 0.35 | 272 | 260 (95.6%) | 94.90% | 0.3 | 0.83 | < 0.001 |
| LAVI, ml/m<sup>2</sup> | 26.6 ± 11.3 | 26.6 ± 11.3 | 289 | 246 (85.1%) | 98.60% | 0.7 | > 0.99 | < 0.001 |
| AV Vmax, m/s | 1.42 ± 0.39 | 1.41 ± 0.38 | 317 | 313 (98.7%) | 97.80% | 0.2 | 0.95 | < 0.001 |
| AV mean PG, mmHg | 5.0 ± 3.3 | 4.8 ± 3.2 | 317 | 313 (98.7%) | 98.70% | 1.6 | 0.96 | < 0.001 |
| GLS***, % | 19.7 ± 3.4 | NA | 296 | 285 (96.3%) | NA | NA | NA | NA |
| TAPSE, mm | 21.4 ± 5.0 | 21.5 ± 4.9 | 136 | 135 (99.3%) | 98.50% | 0.7 | > 0.99 | < 0.001 |

\* Data were considered acceptable if the difference between AI's initial measurements and the final report values were within acceptable ranges listed in Supplemental Table 4.
\*\* The average absolute modification by the sonographer and echocardiologists during the re-evaluation of the AI's initial analysis. This metric represents the magnitude of adjustments performed by the sonographer to ensure the accuracy of the final report.
\*\*\* GLS was not routinely included in clinical reports
IVSTd, Interventricular septal thickness in diastole; PWTd, Posterior wall thickness in diastole; LVIDd, Left ventricular internal diameter in diastole; LVIDs, Left ventricular internal diameter in systole; LVEDV, Left ventricular end-diastolic volume; LVESV, Left ventricular end-systolic volume; LVEF (2D disk), Left ventricular ejection fraction (2D disk method); MV-E, Mitral valve early diastolic velocity; MV-A, Mitral valve late diastolic velocity; E/A, Early to late ventricular filling velocity ratio; DcT, Deceleration time; e' (sep), Septal mitral annular velocity; e' (lat), Lateral mitral annular velocity; E/e' (sep), Ratio of early diastolic velocity to septal mitral annular velocity; E/e' (lat), Ratio of early diastolic velocity to lateral mitral annular velocity; TR Vmax, Maximum tricuspid regurgitation velocity; LAVI, Left atrial volume index; AV Vmax, Maximum aortic valve velocity; AV mean PG, Mean aortic valve pressure gradient; GLS, Global longitudinal strain; TAPSE, Tricuspid annular plane systolic excursion.

Once the images were analyzed by the AI, sonographers and echocardiologists reviewed the parameters and adjusted them for the final report if necessary. Notably, AI’s initial measurements were deemed acceptable for clinical use without significant modification in over 90% of cases for nearly all parameters, with the exception of LV internal diameter (in diastole: 86.7%, in systole: 70.6%; definitions of the acceptable ranges are summarized in Supplemental Table 4). The mean absolute modification by sonographers, calculated as the sum of the absolute differences between AI’s initial measurements and the final report values divided by the number of modified AI values, showed that the modifications required were small, even when necessary. For example, the mean absolute modification was 3.9% for LV ejection fraction and 3.4% for LV end-diastolic diameter.

Scatterplots and Bland-Altman plots were used to visualize the systematic errors between AI’s initial measurements and the final report values, as shown in Figure 3 and Supplemental Figures 2. ICCs between the AI and sonographers exceeded 0.8 for all parameters as shown in Table 2, with p-values less than 0.001, indicating a high level of agreement. For systolic and diastolic LV diameters and wall thicknesses, it was observed that AI tended to return more extreme values—i.e., larger values were slightly overestimated and smaller values were slightly underestimated.

**Figure 3.**
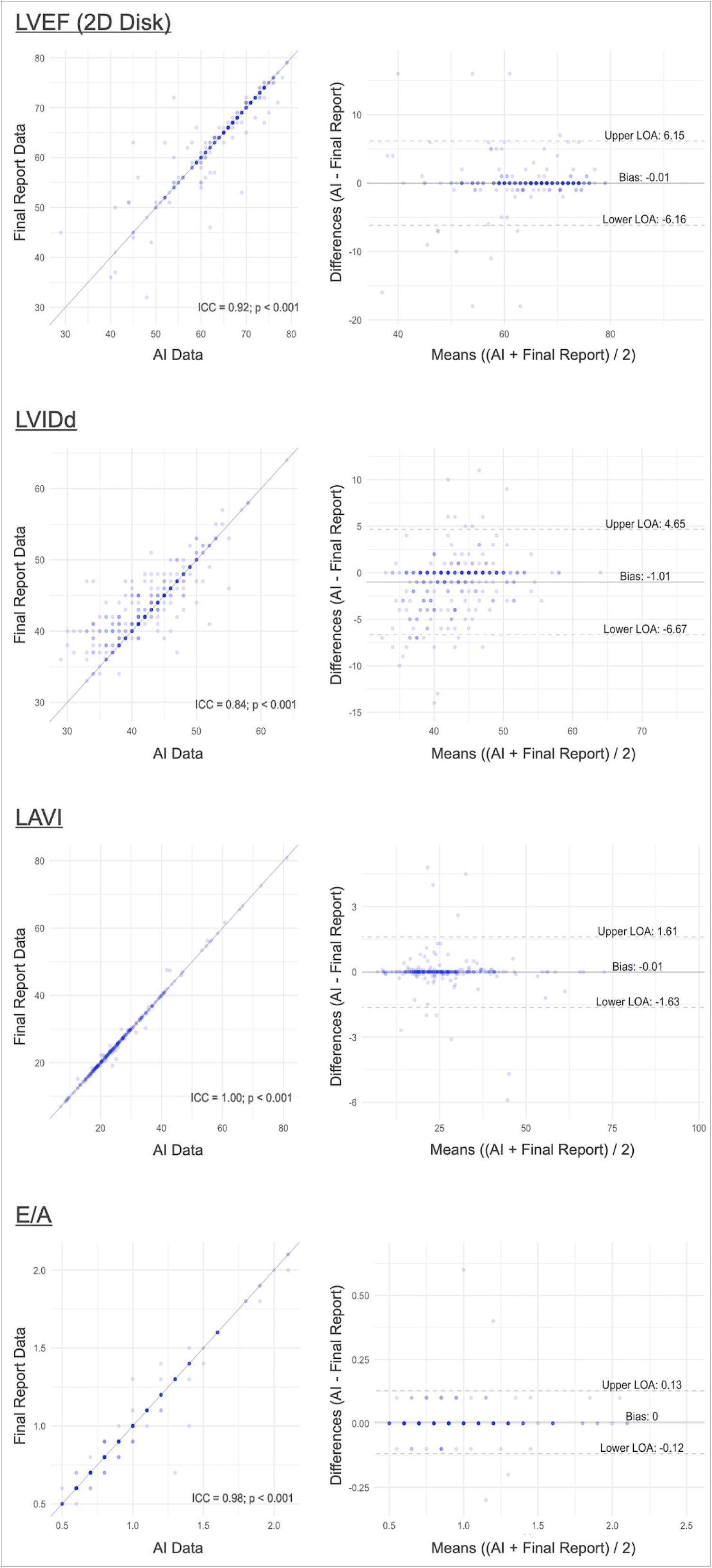
Concordance of AI’s Initial Measurements and the Final Values The scatter plots (left) demonstrate a strong correlation between AI-generated data and the final report values. The Bland-Altman plots illustrate the mean differences (bias) and limits of agreement (LOA) between the two measurement sets, with bias close to zero for each parameter. The overall concordance was excellent, with ICCs exceeding 0.8 and narrow LOAs.

## Discussions

This first-ever trial to randomize the use of an AI-based automated tool on a daily basis revealed that AI significantly enhanced the efficiency of screening echocardiography, reducing examination time despite a 3.4-fold increase in the number of parameters measured. This improved efficiency increased the number of examinations per day without increasing sonographers’ fatigue; in fact, it mitigated mental fatigue. Furthermore, being freed from the need to perform time-consuming measurements allowed sonographers to focus on image acquisition, which led to improved image quality.

AI-based tools in echocardiography have been extensively investigated since approximately 2018 ^20, 21^, with numerous studies demonstrating their accuracy and efficiency in analyzing echocardiographic images. For example, a retrospective study using the same AI tool employed in our trial confirmed its capacity to accurately analyze echocardiographic parameters under controlled conditions ^11^. Another experimental study reported a 70% reduction in analysis and reporting time with this AI tool ^9^. Despite these promising results, AI has not yet achieved widespread adoption in clinical practice. A significant barrier to broader implementation is that many studies evaluating AI tools, including those mentioned above, were conducted in retrospective or experimental settings, which do not fully capture the operational complexities of routine clinical workflows.

Recently, several studies have begun addressing this gap by evaluating AI tools in prospective, randomized trials ^22, 23^. In contrast to retrospective studies, which lack the complexity of real-world clinical settings, these randomized trials provide more practical insights. One notable example is EchoNet-RCT, the first randomized controlled trial to evaluate AI in echocardiography, which demonstrated the precision of AI in automatically analyzing echocardiographic images to measure LV ejection fraction and validated the accuracy of AI-guided workflows in clinical practice ^8^. Our study, utilizing an AI tool capable of analyzing a broad range of clinical echocardiographic parameters, extends this focus beyond AI accuracy to examine its broader clinical impact.

Specifically, our findings demonstrated that integrating AI into daily echocardiography workflows significantly enhanced efficiency by reducing total examination time, while simultaneously increasing the number of examinations performed per day. On AI days, sonographers were able to complete more examinations without sacrificing diagnostic quality. The AI tool not only automated the analysis of routine echocardiographic parameters but also facilitated the acquisition of more comprehensive data, including complex measurements like LV strain, which are not typically analyzed during routine screening. Importantly, the diagnostic integrity of these AI-generated measurements was maintained, as evidenced by the high concordance between AI’s initial measurements and the final expert-reviewed reports. This consistency underscores AI’s potential to augment sonographers’ capacity for routine echocardiographic assessments, allowing for increased throughput without compromising clinical accuracy.

A critical finding of our study was the potential for AI to alleviate the mental and physical burden on sonographers, as indicated by the questionnaire results—an especially relevant benefit in high-volume echocardiographic laboratories. Screening echocardiography, often viewed as repetitive and routine, requires sonographers to conduct rapid yet accurate assessments. These repetitive tasks, combined with increasing clinical demands, may reduce motivation and increase fatigue among sonographers ^2^. Our results suggest that AI can help mitigate these challenges. Moreover, while the time saved through AI-enhanced efficiency in our study was used to perform more examinations, this additional time could be repurposed for more complex clinical activities, such as discussing hemodynamic findings or treatment strategies with clinicians, or providing more detailed explanations to patients. Engaging in these more intellectually stimulating and patient-centered tasks could enhance job satisfaction for sonographers, contributing to a more rewarding and sustainable clinical practice.

Another significant outcome from our trial was the improvement in the quality of echocardiographic images on AI-assisted days. High-quality imaging is crucial for ensuring diagnostic accuracy, particularly for clinicians reviewing the images after acquisition. It is likely that the observed improvement in image quality on AI days occurred because sonographers were able to devote more attention to image acquisition when relieved of the cognitive load of performing manual measurements. Additionally, sonographers likely recognized that AI algorithms perform optimally with high-quality images, motivating them to ensure the best possible image acquisition under AI guidance, even in cases where they might have previously accepted suboptimal images when manually handling measurements. This enhancement in image quality not only facilitates better AI performance but also ensures that subsequent clinical decisions are based on clearer, more accurate data, further underscoring the value of integrating AI into routine echocardiographic practice.

Looking ahead, the integration of AI into echocardiographic practice presents a promising opportunity to enhance workflow efficiency, reduce sonographers’ burden, and potentially support more personalized and comprehensive patient care. Further refinement of AI tools and larger-scale studies will be necessary to fully assess their clinical utility and pave the way for broader adoption in routine practice.

### Limitations

This study has several limitations. First, it was conducted at a single center, and since echocardiography protocols can differ across institutions, multi-center studies are needed to confirm the generalizability of these results. Additionally, the study duration was limited to about two and a half months, so it remains unclear whether the observed efficiency gains, workload improvements, and enhancements in image quality would persist over a longer period. Second, although the trial was randomized, sonographers were aware of AI usage, which could have influenced their performance, even though they were blinded to the study’s endpoints. This awareness may have introduced some bias. Third, this study focused on experienced sonographers, leaving open the question of whether similar improvements in time efficiency and outcomes would be observed with less experienced operators. Prior research suggests that AI may enhance performance even for novices ^24^, but fundamental sonography skills might still be required to fully leverage the benefits of AI. Next, our study focused on workflow efficiency and diagnostic quality but did not directly assess how AI-assisted echocardiography impacts patient outcomes or clinical decision-making. Lastly, the majority of patients included were undergoing screening echocardiography, primarily in sinus rhythm and with low rates of complex cardiac pathology. This may limit the applicability of our findings to more complex clinical cases, such as those with advanced heart failure or significant valvular disease, where human oversight might be more critical. Further research is needed to assess the impact of AI on both workflow efficiency and clinical outcomes in a broader range of clinical scenarios.

## Conclusions

This randomized trial demonstrated that integrating an AI-based automated analysis system into clinical echocardiography workflows can significantly reduce examination time and increase the number of examinations performed per day, thereby enhancing the overall efficiency of the echocardiography laboratory. Importantly, AI analysis maintained diagnostic quality, mitigated sonographers’ fatigue, and improved image quality, supporting its potential role in streamlining clinical practice. Continued advancements in AI technology hold promise for further improvements in both workflow efficiency and diagnostic accuracy in echocardiography.

## Contributors

NK and TM supervised the study. NK designed the study. AS, NK, ES, YN, AM, TK, and SM conducted the study. AS and NK performed the statistical analyses. AS and ES assessed image quality. YA and KS coordinated the implementation of the AI analysis software. All authors had access to the data presented in the study, contributed to critical revisions, and approved the final version of the manuscript. All authors take responsibility for the integrity of the work and were responsible for the decision to submit for publication.

## Data sharing statement

The data underlying this article cannot be shared publicly for the reason of maintaining the privacy of individuals who participated in the study. The data will be shared on reasonable request to the corresponding author.

## Trial registration name and number

Artificial Intelligence-based automated ECHOcardiographic measurements and the workflow of sonographers: Randomized Control Trial (AI-ECHO RCT) UMIN000053259

## Acknowledgements

This work was supported by the Japan Society for the Promotion of Science KAKENHI (grant number 21K18086). We thank the ethics committee of the Faculty of Medicine, Juntendo University, for their approval and oversight. We also acknowledge M3AI Inc. for providing technical support related to the AI analysis software and thank the clinical staff and patients who participated in this study.

## Abbreviation

AI: artificial intelligence
ICC: intraclass correlation coefficients
LV: left ventricular

