## Supplemental Table for "Artificial Intelligence-based Automated Echocardiographic Analysis and the Workflow of Sonographers: A randomized crossover trial"

**Supplemental Tables**

### Supplemental Table 1. CONSORT 2010 checklist of information to include when reporting a randomized trial

| Section/Topic | Item No | Checklist item | Reported on page No |
| --- | --- | --- | --- |
| Title and abstract | | | |
|  | 1a | Identification as a randomized trial in the title | Page 1 |
|  | 1b | Structured summary of trial design, methods, results, and conclusions (for specific guidance see CONSORT for abstracts) | Page 2-3 |
| Introduction | | | |
| Background and objectives | 2a | Scientific background and explanation of rationale | Page 6 |
|  | 2b | Specific objectives or hypotheses | Page 6 |
| Methods | | | |
| Trial design | 3a | Description of trial design (such as parallel, factorial) including allocation ratio | Page 7 |
|  | 3b | Important changes to methods after trial commencement (such as eligibility criteria), with reasons | N/A |
| Participants | 4a | Eligibility criteria for participants | Page 7-8 |
|  | 4b | Settings and locations where the data were collected | Page 7-8 |
| Interventions | 5 | The interventions for each group with sufficient details to allow replication, including how and when they were actually administered | Page 7-8 |
| Outcomes | 6a | Completely defined pre-specified primary and secondary outcome measures, including how and when they were assessed | Page 8-9 |
|  | 6b | Any changes to trial outcomes after the trial commenced, with reasons | N/A |
| Sample size | 7a | How sample size was determined | Page 9 |
|  | 7b | When applicable, explanation of any interim analyses and stopping guidelines | N/A |
| Randomization: |  |  |  |
| Sequence generation | 8a | Method used to generate the random allocation sequence | Page 7-8 |
|  | 8b | Type of randomization; details of any restriction (such as blocking and block size) | Page 7-8 |
| Allocation concealment mechanism | 9 | Mechanism used to implement the random allocation sequence (such as sequentially numbered containers), describing any steps taken to conceal the sequence until interventions were assigned | Page 7-8 |
| Implementation | 10 | Who generated the random allocation sequence, who enrolled participants, and who assigned participants to interventions | Page 7-8 |
| Blinding | 11a | If done, who was blinded after assignment to interventions (for example, participants, care providers, those assessing outcomes) and how | N/A |
|  | 11b | If relevant, description of the similarity of interventions | N/A |
| Statistical methods | 12a | Statistical methods used to compare groups for primary and secondary outcomes | Page 9-10 |
|  | 12b | Methods for additional analyses, such as subgroup analyses and adjusted analyses | N/A |
| Results | | | |
| Participant flow (a diagram is strongly recommended) | 13a | For each group, the numbers of participants who were randomly assigned, received intended treatment, and were analyzed for the primary outcome | Page 10 |
|  | 13b | For each group, losses and exclusions after randomization, together with reasons | N/A |
| Recruitment | 14a | Dates defining the periods of recruitment and follow-up | Page 10 |
|  | 14b | Why the trial ended or was stopped | N/A |
| Baseline data | 15 | A table showing baseline demographic and clinical characteristics for each group | Page 10,  Table 1 |
| Numbers analyzed | 16 | For each group, number of participants (denominator) included in each analysis and whether the analysis was by original assigned groups | Page 10,  Table 1 |
| Outcomes and estimation | 17a | For each primary and secondary outcome, results for each group, and the estimated effect size and its precision (such as 95% confidence interval) | Table 2,  Figure 1, 2, 3  Page 10-12 |
|  | 17b | For binary outcomes, presentation of both absolute and relative effect sizes is recommended | N/A |
| Ancillary analyses | 18 | Results of any other analyses performed, including subgroup analyses and adjusted analyses, distinguishing pre-specified from exploratory | N/A |
| Harms | 19 | All important harms or unintended effects in each group (for specific guidance see CONSORT for harms) | N/A |
| Discussion | | | |
| Limitations | 20 | Trial limitations, addressing sources of potential bias, imprecision, and, if relevant, multiplicity of analyses | Page 14-15 |
| Generalizability | 21 | Generalizability (external validity, applicability) of the trial findings | Page 12-14 |
| Interpretation | 22 | Interpretation consistent with results, balancing benefits and harms, and considering other relevant evidence | Page 12-14 |
| Other information | | |  |
| Registration | 23 | Registration number and name of trial registry | Page 5 |
| Protocol | 24 | Where the full trial protocol can be accessed, if available | Page 1 |
| Funding | 25 | Sources of funding and other support (such as supply of drugs), role of funders | Page 4 |

### Supplemental Table 2. Percentage of Echocardiographic Parameters Collected on AI and Non-AI Days

|  | **Non-AI days (%)** | **AI days (%)** |
| --- | --- | --- |
| IVSTd | 100 | 100 |
| PWTd | 100 | 100 |
| LVIDd | 100 | 100 |
| LVIDs | 100 | 100 |
| LA diameter | 100 | 100 |
| Ascending Aorta | 98.5 | 97.8 |
| IVC (inspire) | 100 | 99.4 |
| IVC (expire) | 96.3 | 87.7 |
| LVEF (Teichholz) | 100 | 100 |
| Fractional Shortening | 100 | 100 |
| LVEDV | 93.3 | 86.8 |
| LVESV | 93.3 | 86.8 |
| LVEF (2D disk) | 93.3 | 86.8 |
| AV Vmax | 100 | 100 |
| MV-E | 100 | 100 |
| MV-A | 95.9 | 97.2 |
| E/A | 95.9 | 96.8 |
| DcT | 100 | 100 |
| e' (sep) | 100 | 100 |
| e (lat) | 100 | 100 |
| E/e (sep) | 100 | 99.7 |
| E/e (lat) | 100 | 99.7 |
| TR Vmax | 92.5 | 88.3 |
| Estimated RVSP | 92.5 | 86.8 |
| Estimated RAP | 100 | 100 |
| LVOT diameter | 13.1 | 93.4 |
| RVOT diameter | 2.6 | 0.3 |
| AV mean PG | 5.6 | 100 |
| AVA (2D) | 3.7 | 3.2 |
| AVA (Doppler) | 4.9 | 64.0 |
| AVA index (Doppler) | 4.9 | 63.4 |
| RWT | 0 | 97.5 |
| LV length (A2C) | 0 | 95.6 |
| LV length (A4C) | 0 | 96.2 |
| LV mass | 0 | 97.2 |
| LV mass index | 0 | 96.2 |
| LVEDV (2D disk, A2C) | 0 | 90.9 |
| LVEDV (2D disk, A4C) | 0 | 95.3 |
| LVEDV index (2D disk, A2C) | 0 | 89.9 |
| LVEDV index (2D disk, A4C) | 0 | 94.6 |
| LVEDV index (2D disk, biplane) | 0 | 87.4 |
| LVESV (2D disk, A2C) | 0 | 90.9 |
| LVESV (2D disk, A4C) | 0 | 95.3 |
| LVESV index (2D disk, A2C) | 0 | 89.9 |
| LVESV index (2D disk, A4C) | 0 | 94.6 |
| LVESV index (2D disk, biplane) | 0 | 87.4 |
| LVEF (2D disk, A2C) | 0 | 90.9 |
| LVEF (2D disk, A4C) | 0 | 95.3 |
| LV Stroke Volume (2D disk, biplane) | 0 | 88.0 |
| LV Cardiac Output (2D disk, biplane) | 0 | 88.0 |
| LVOT max PG | 0 | 99.4 |
| LVOT mean PG | 0 | 99.4 |
| LVOT Vmax | 0 | 99.4 |
| LVOT Vmean | 0 | 99.4 |
| LVOT VTI | 0 | 99.4 |
| AV Vmean | 0 | 98.7 |
| AV max PG | 0 | 98.7 |
| AV VTI | 0 | 98.7 |
| GLS | 0 | 89.9 |
| GLS (A2C) | 0 | 95.0 |
| GLS (A3C) | 0 | 94.6 |
| GLS (A4C) | 0 | 96.2 |
| LA length (A2C) | 0 | 87.4 |
| LA length (A4C) | 0 | 91.8 |
| LA Reservoir | 0 | 65.0 |
| LA Reservoir (A2C) | 0 | 71.9 |
| LA Reservoir (A4C) | 0 | 80.1 |
| LA width (Diastolic A2C) | 0 | 87.4 |
| LA width (Systolic A4C) | 0 | 91.8 |
| LAESV (2D disk, A2C) | 0 | 82.3 |
| LAESV (2D disk, A4C) | 0 | 88.6 |
| LAESV (2D disk, biplane) | 0 | 77.6 |
| LAESV index (2D disk, A2C) | 0 | 81.4 |
| LAESV index (2D disk, A4C) | 0 | 88.0 |
| LAESV index (2D disk, biplane) | 0 | 74.1 |
| e' (mean) | 0 | 95.3 |
| E/e' (mean) | 0 | 79.5 |
| a' (lat) | 0 | 97.2 |
| a' (sep) | 0 | 93.7 |
| s' (lat) | 0 | 99.7 |
| s' (sep) | 0 | 97.5 |
| RVIDd (basal) | 0 | 79.2 |
| RVIDd (mid) | 0 | 68.8 |
| RV area (Diastolic A4C) | 0 | 65.9 |
| RV area (Systolic A4C) | 0 | 65.9 |
| RV FAC | 0 | 64.7 |
| RVEDV (2D disk, A4C) | 0 | 86.4 |
| RVESV (2D disk, A4C) | 0 | 75.1 |
| RV a' | 0 | 42.6 |
| RV e' | 0 | 44.8 |
| RV s' | 0 | 44.8 |
| RV/LV ratio | 0 | 78.9 |
| RA length (A4C) | 0 | 81.4 |
| RA width (A4C) | 0 | 81.4 |
| RA area (A4C) | 0 | 81.4 |
| RAESV (2D disk, A4C) | 0 | 81.4 |
| TAPSE | 0 | 42.6 |
| TR max PG | 0 | 82.0 |
| PASP | 0 | 72.6 |

IVSTd, Interventricular septal thickness in diastole; PWTd, Posterior wall thickness in diastole; LVIDd, Left ventricular internal diameter in diastole; LVIDs, Left ventricular internal diameter in systole; LA diameter, Left atrial diameter; Ascending Aorta, Ascending aorta diameter; IVC, Inferior vena cava diameter; LVEF, Left ventricular ejection fraction; LVEDV, Left ventricular end-diastolic volume; LVESV, Left ventricular end-systolic volume; AV Vmax, Aortic valve maximum velocity; MV-E, Mitral valve early diastolic velocity; MV-A, Mitral valve late diastolic velocity; E/A, Early to late ventricular filling velocity ratio; DcT, Deceleration time; e', Early diastolic velocity; E/e', Ratio of E to e'; TR Vmax, Tricuspid regurgitation maximum velocity; RVSP, Right ventricular systolic pressure; RAP, Right atrial pressure; LVOT, Left ventricular outflow tract; RVOT, Right ventricular outflow tract; AVA, Aortic valve area; RWT, Relative wall thickness; LV mass, Left ventricular mass; GLS, Global longitudinal strain; LAESV, Left atrial end-systolic volume; RVIDd, Right ventricular internal diameter; RV area, Right ventricular area; RV FAC, Right ventricular fractional area change; RV a', Right ventricular late diastolic velocity; RV e', Right ventricular early diastolic velocity; RV s', Right ventricular systolic velocity; RV/LV ratio, Right ventricular/Left ventricular ratio; RA length (A4C), Right atrial length in apical 4-chamber view; RA width (A4C), Right atrial width in apical 4-chamber view; RA area (A4C), Right atrial area in apical 4-chamber view; RAESV, Right atrial end-systolic volume; TAPSE, Tricuspid annular plane systolic excursion; TR max PG, Tricuspid regurgitation maximum pressure gradient; PASP, Pulmonary artery systolic pressure.

### Supplemental Table 3. Detail of a Daily Questionnaire for sonographers’ mental and physical fatigue at the end of each workday

Please evaluate the mental fatigue you experienced during today's echocardiography tasks.

1 - Do not feel at all
2 - Feel slightly
3 - Neutral
4 - Feel somewhat
5 - Feel strongly

Please evaluate the physical fatigue you experienced during today's echocardiography tasks.

1 - Do not feel at all
2 - Feel slightly
3 - Neutral
4 - Feel somewhat
5 - Feel strongly

Please evaluate the complexity and the time taken for today's echocardiography tasks.

1 - Do not feel at all

2 - Feel slightly

3 - Neutral

4 - Feel somewhat

5 - Feel strongly

### Supplemental Table 4. List of acceptable ranges for differences between the AI's initial measurements and the final report values

| Parameters | Acceptable range |
| --- | --- |
| IVSTd, mm | 2.0 |
| PWTd, mm | 2.0 |
| LVIDd, mm | 4.0 |
| LVIDs, mm | 3.0 |
| LVEDV, ml | 9.0 |
| LVESV, ml | 4.0 |
| LVEF (2D disk), % | 4.0 |
| MV-E, cm/s | 4.0 |
| MV-A, cm/s | 4.0 |
| E/A | 0.20 |
| DcT, ms | 25 |
| e' (sep), cm/s | 0.5 |
| e' (lat), cm/s | 0.5 |
| E/e' (sep) | 0.5 |
| E/e' (lat) | 0.5 |
| TR Vmax, m/s | 0.40 |
| LAVI, ml/m^2^ | 4.0 |
| AV Vmax, m/s | 0.30 |
| AV mean PG, mmHg | 3.0 |
| GLS, % | 3.0 |
| TAPSE, mm | 2.0 |

IVSTd, Interventricular septal thickness in diastole; PWTd, Posterior wall thickness in diastole; LVIDd, Left ventricular internal diameter in diastole; LVIDs, Left ventricular internal diameter in systole; LVEDV, Left ventricular end-diastolic volume; LVESV, Left ventricular end-systolic volume; LVEF (2D disk), Left ventricular ejection fraction (2D disk method); MV-E, Mitral valve early diastolic velocity; MV-A, Mitral valve late diastolic velocity; E/A, Early to late ventricular filling velocity ratio; DcT, Deceleration time; e' (sep), Septal mitral annular velocity; e' (lat), Lateral mitral annular velocity; E/e' (sep), Ratio of early diastolic velocity to septal mitral annular velocity; E/e' (lat), Ratio of early diastolic velocity to lateral mitral annular velocity; TR Vmax, Maximum tricuspid regurgitation velocity; LAVI, Left atrial volume index; AV Vmax, Maximum aortic valve velocity; AV mean PG, Mean aortic valve pressure gradient; GLS, Global longitudinal strain; TAPSE, Tricuspid annular plane systolic excursion.
