## Supplemental Figure for "Artificial Intelligence-based Automated Echocardiographic Analysis and the Workflow of Sonographers: A randomized crossover trial"

**Supplemental Figures**

### Supplemental Figure 1. AI-generated Results with Trace Lines Overlaid on Images

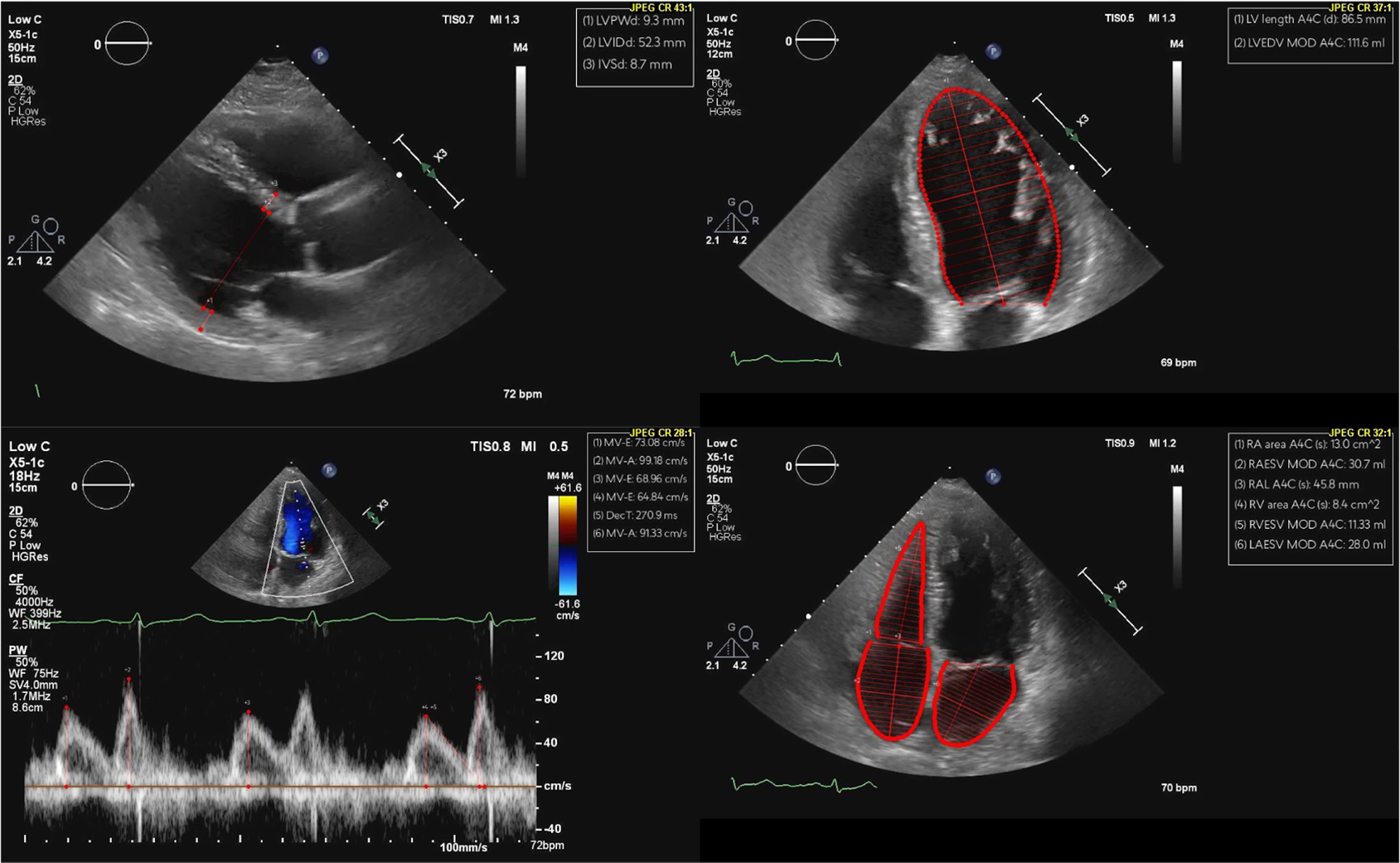

The AI software automatically classifies echocardiographic views and provides measurements, with trace lines overlaid on the images to indicate the AI's analysis.

### Supplemental Figure 2. Concordance of AI’s Initial Measurements and the Final Values

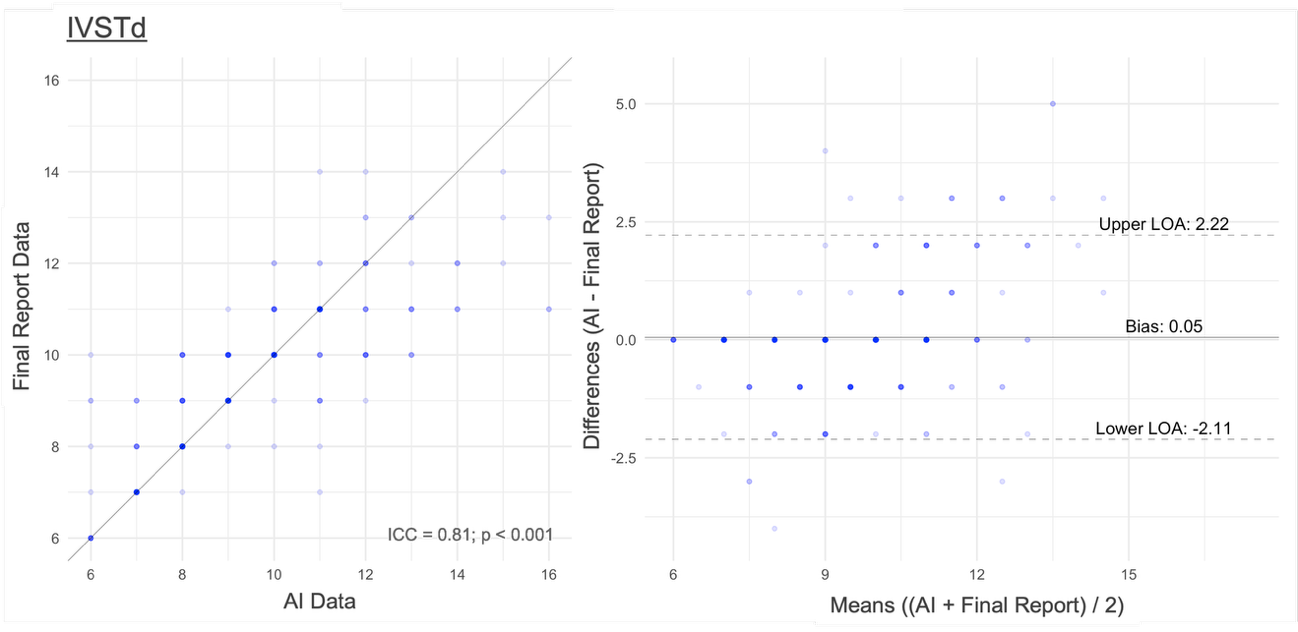

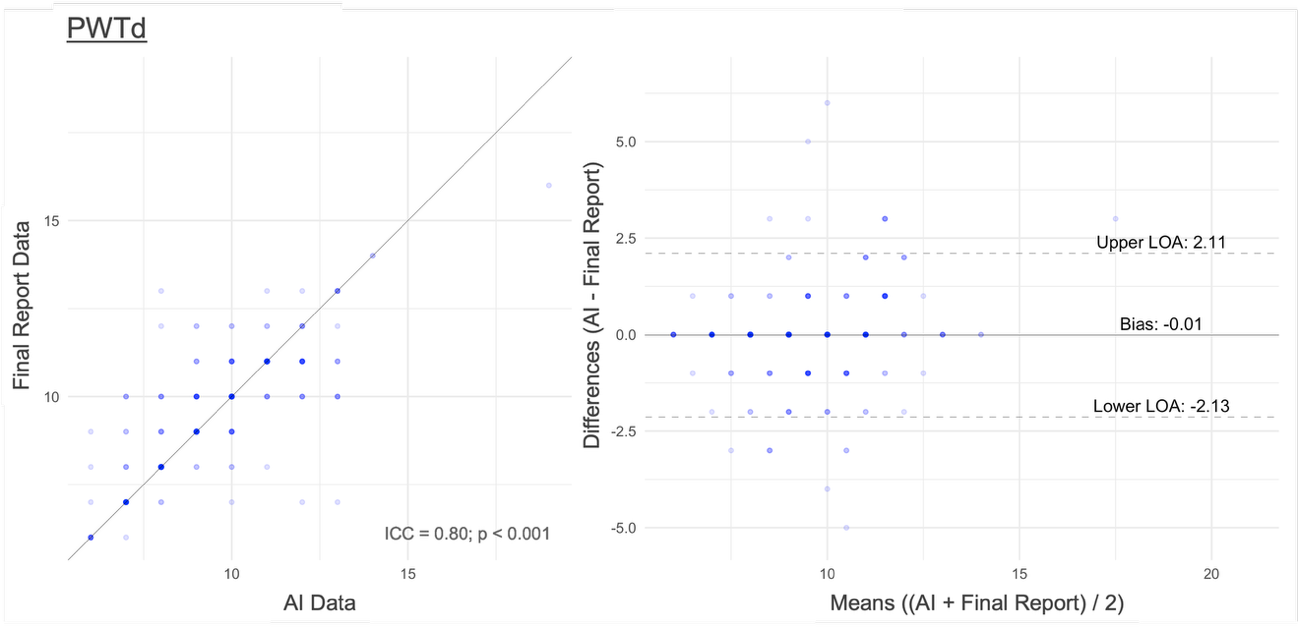

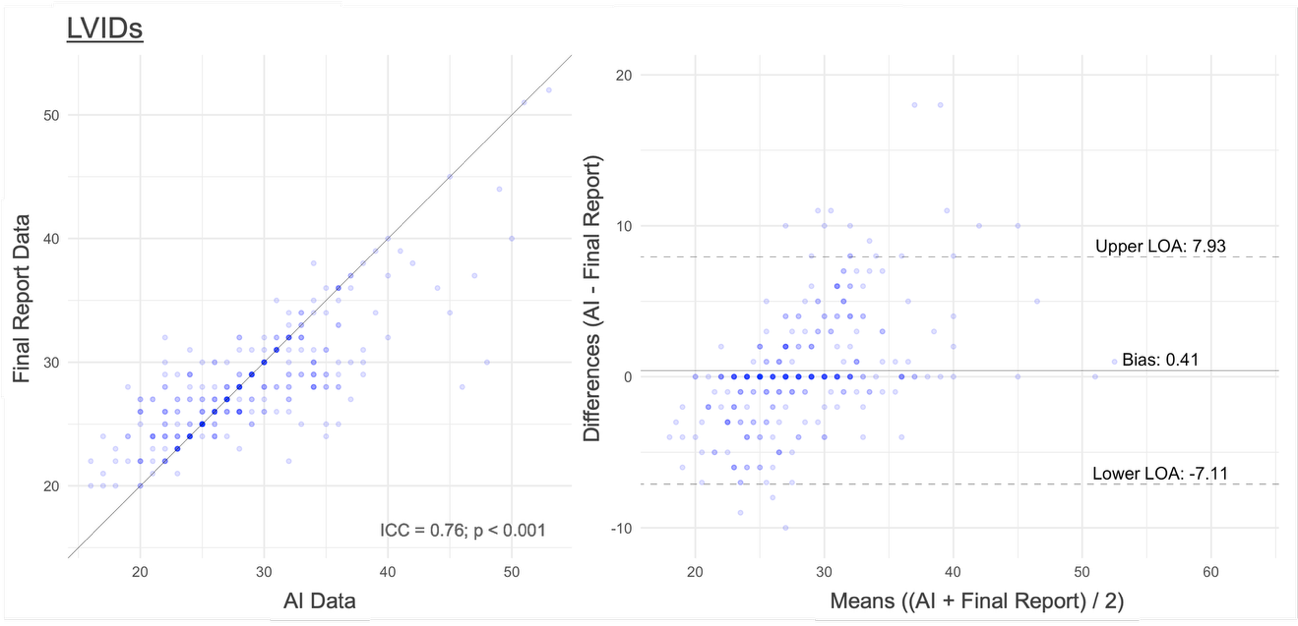

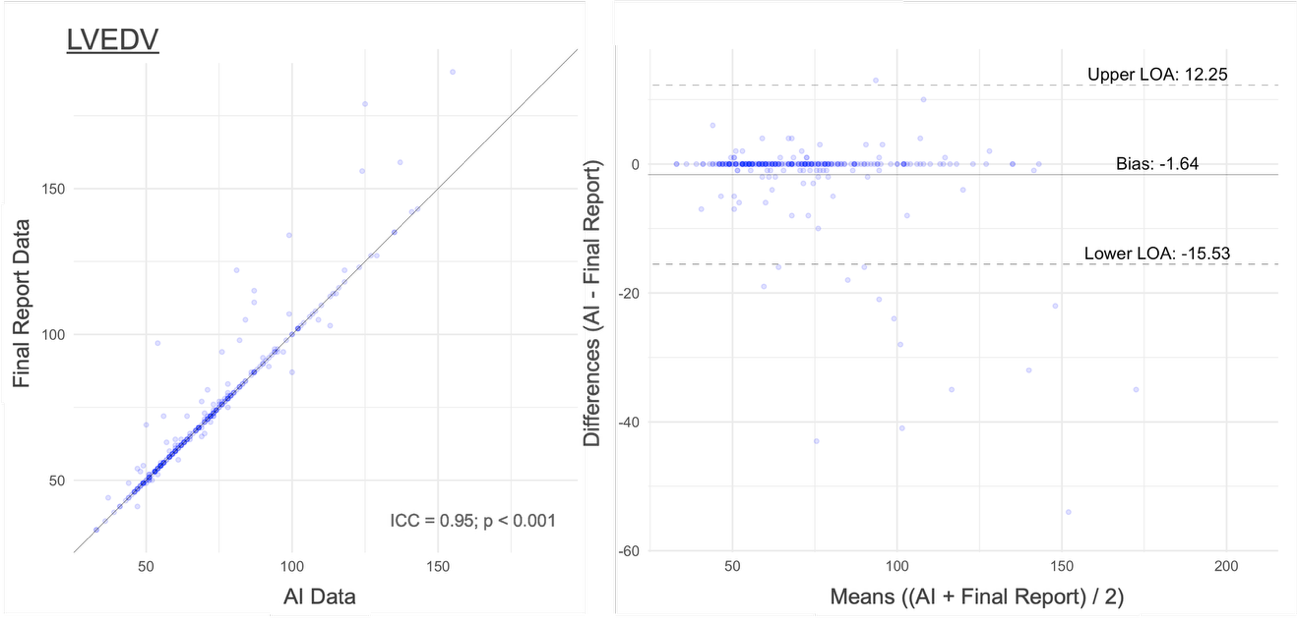

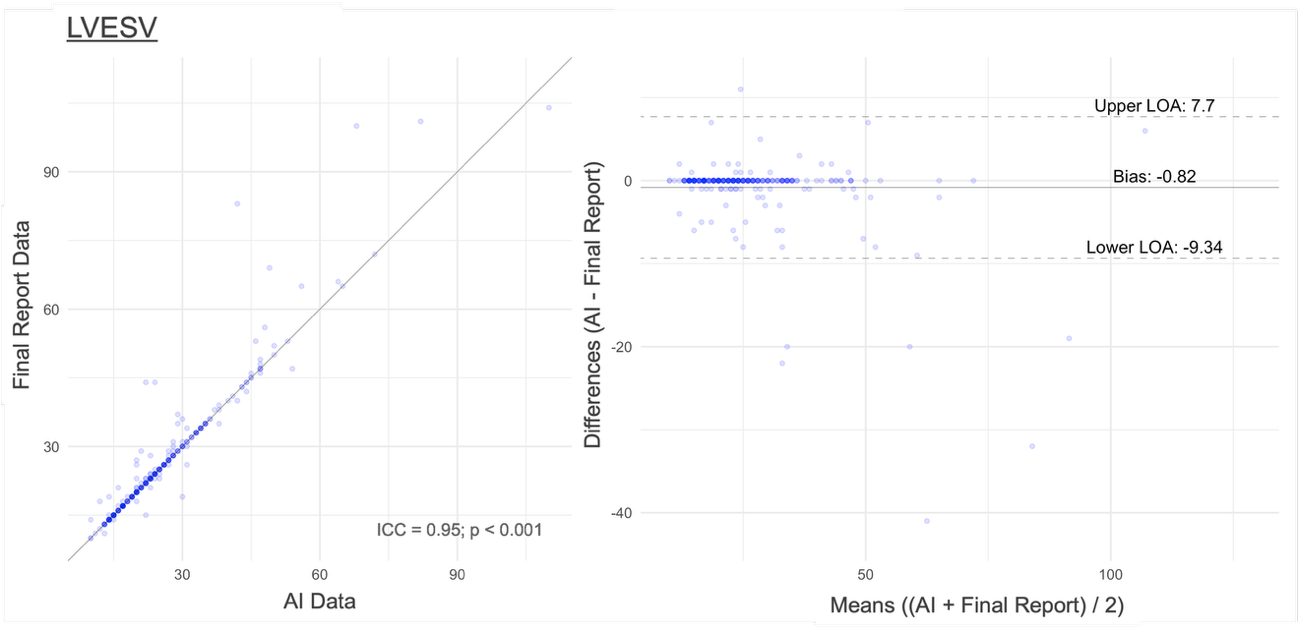

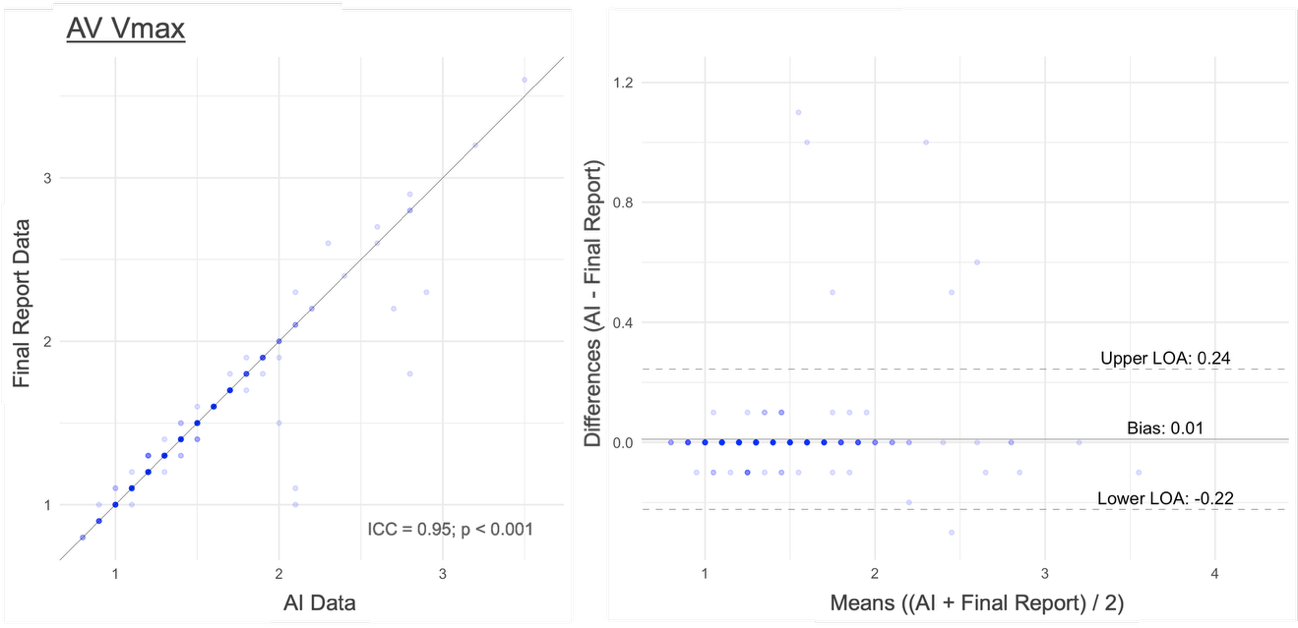

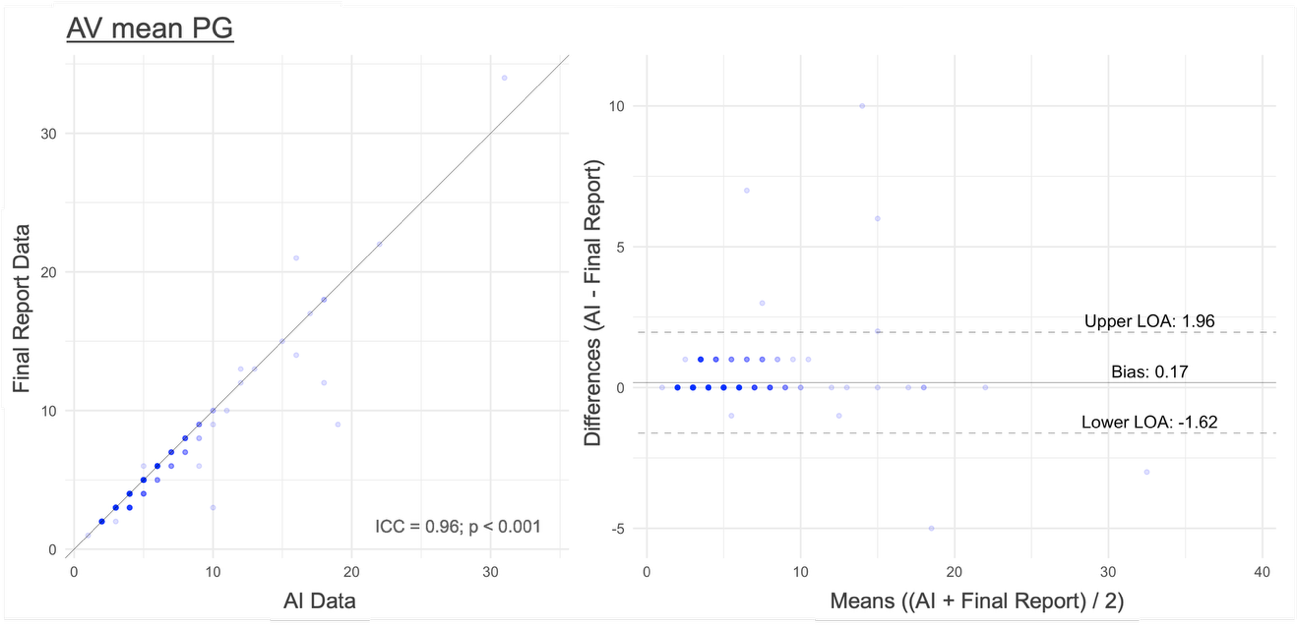

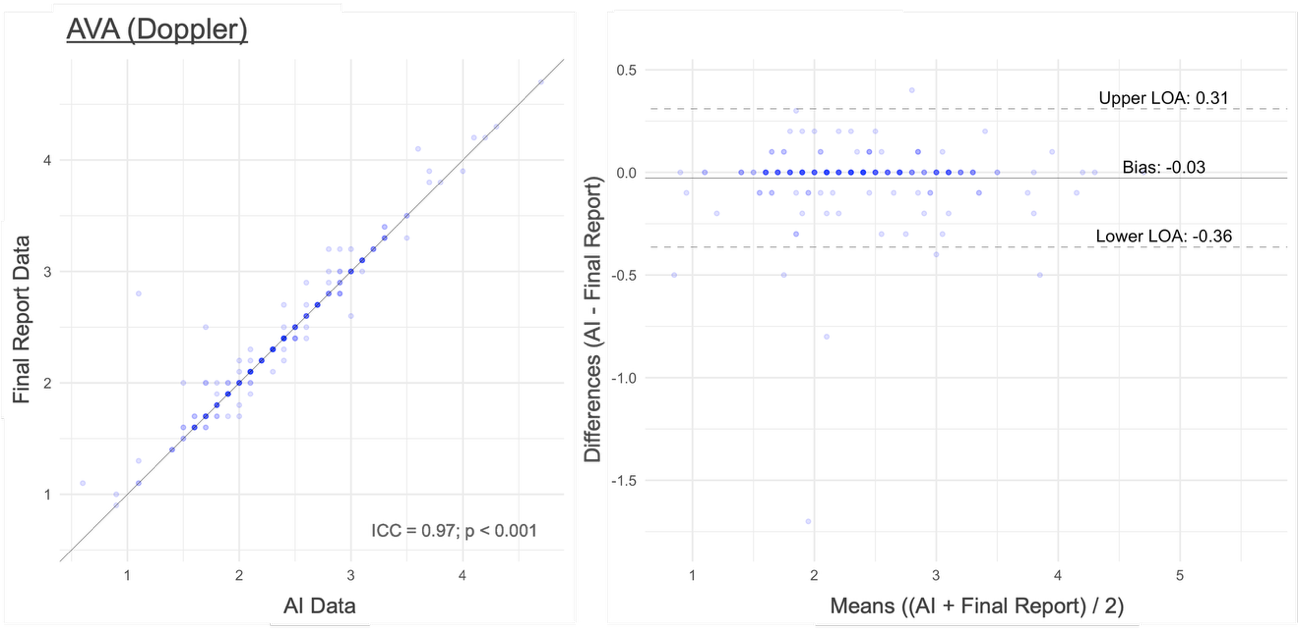

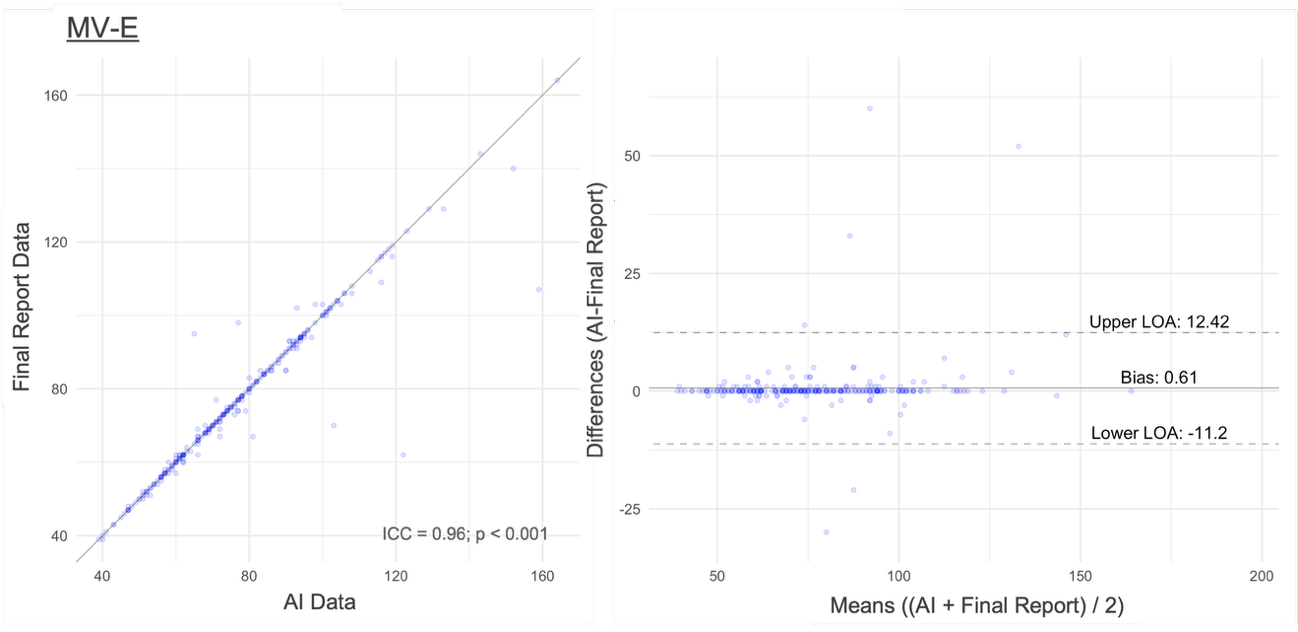

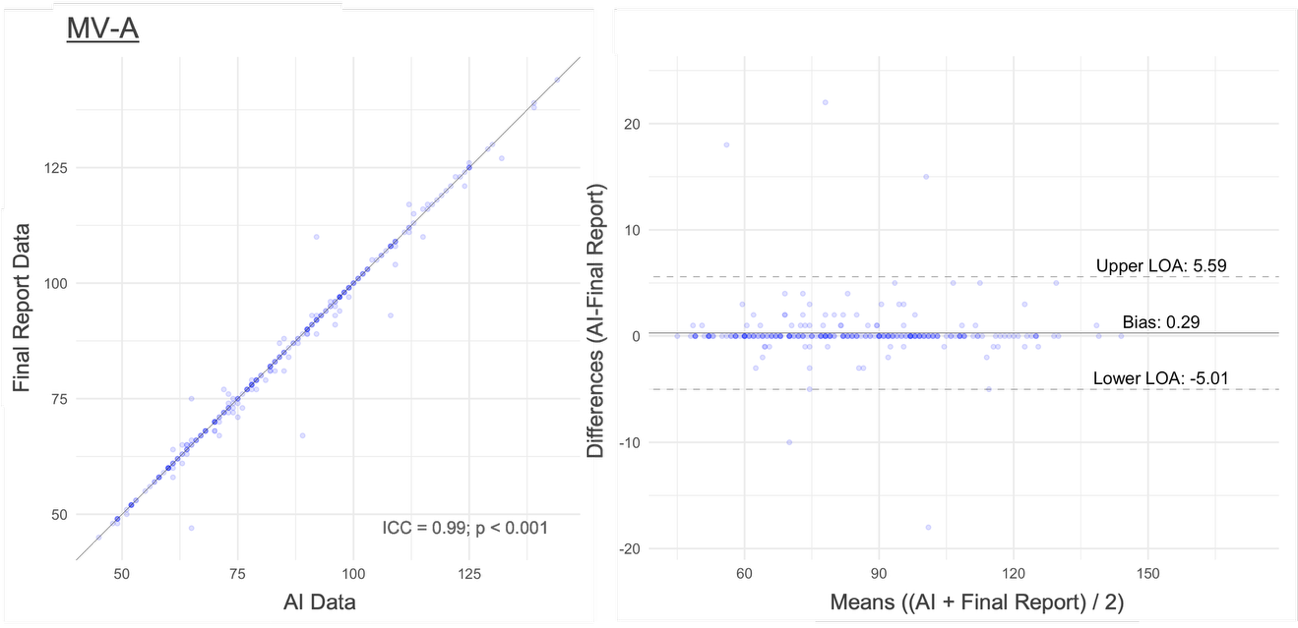

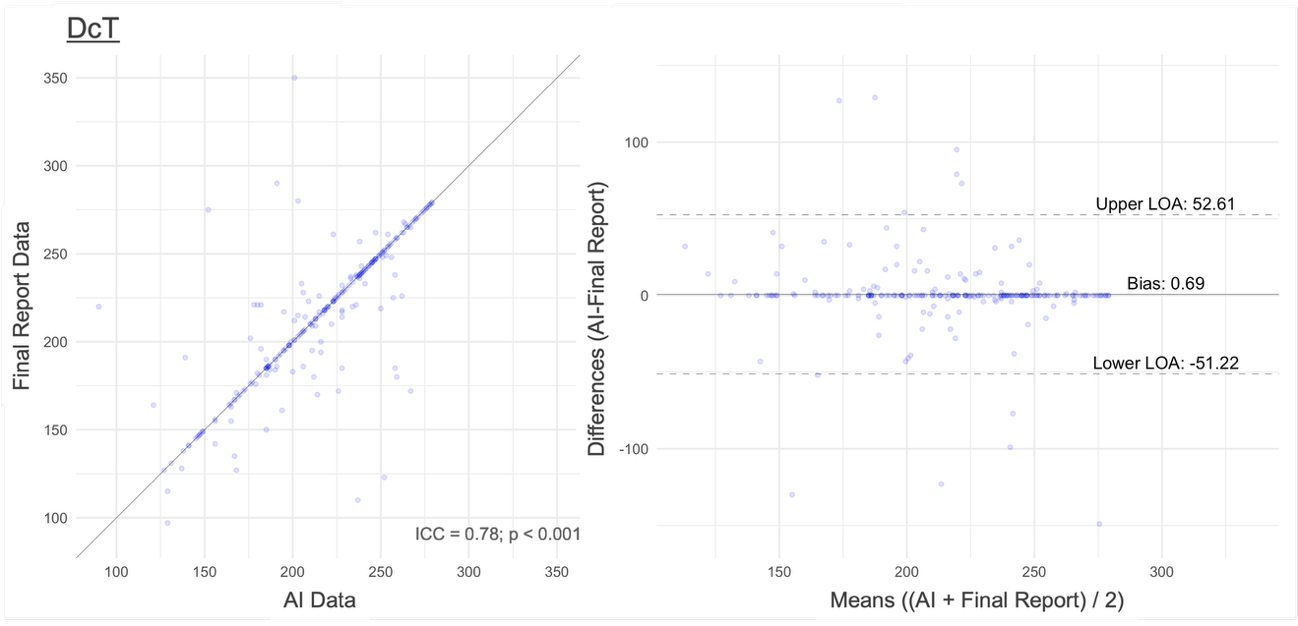

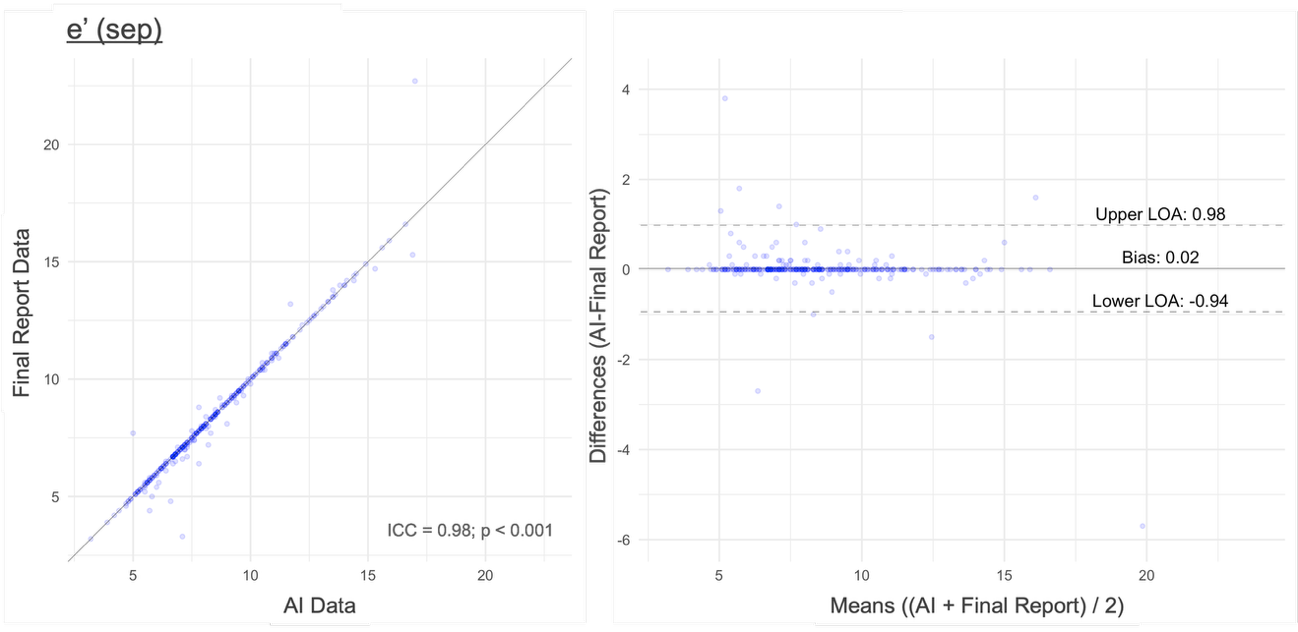

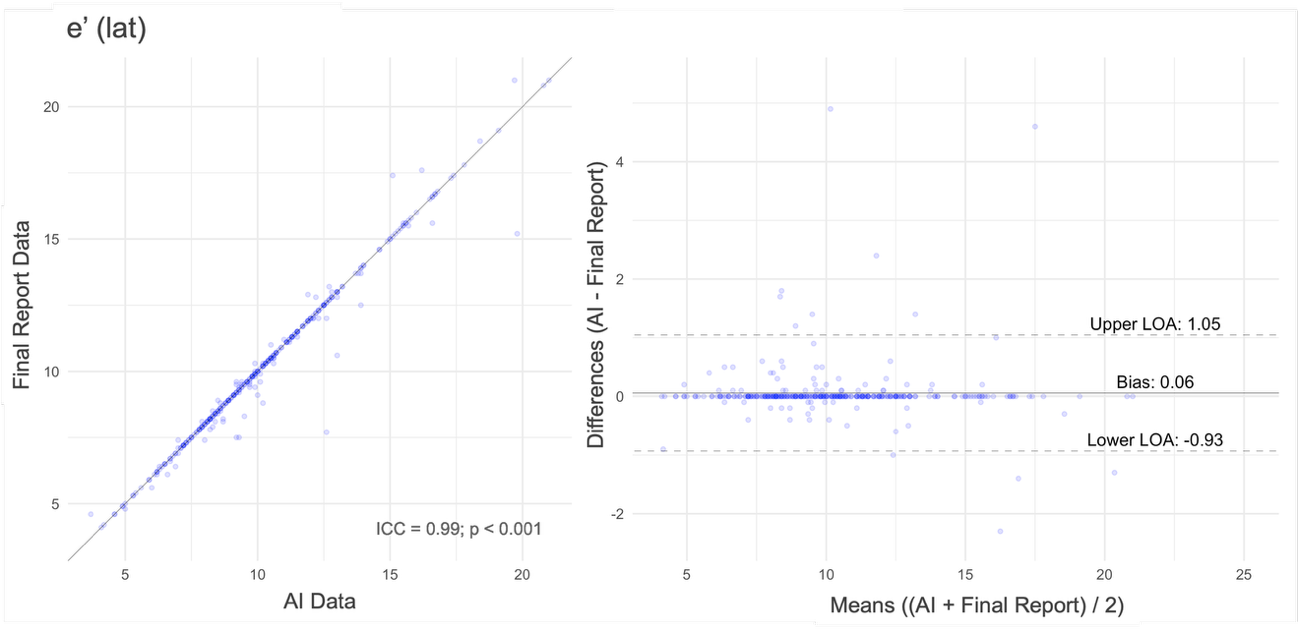

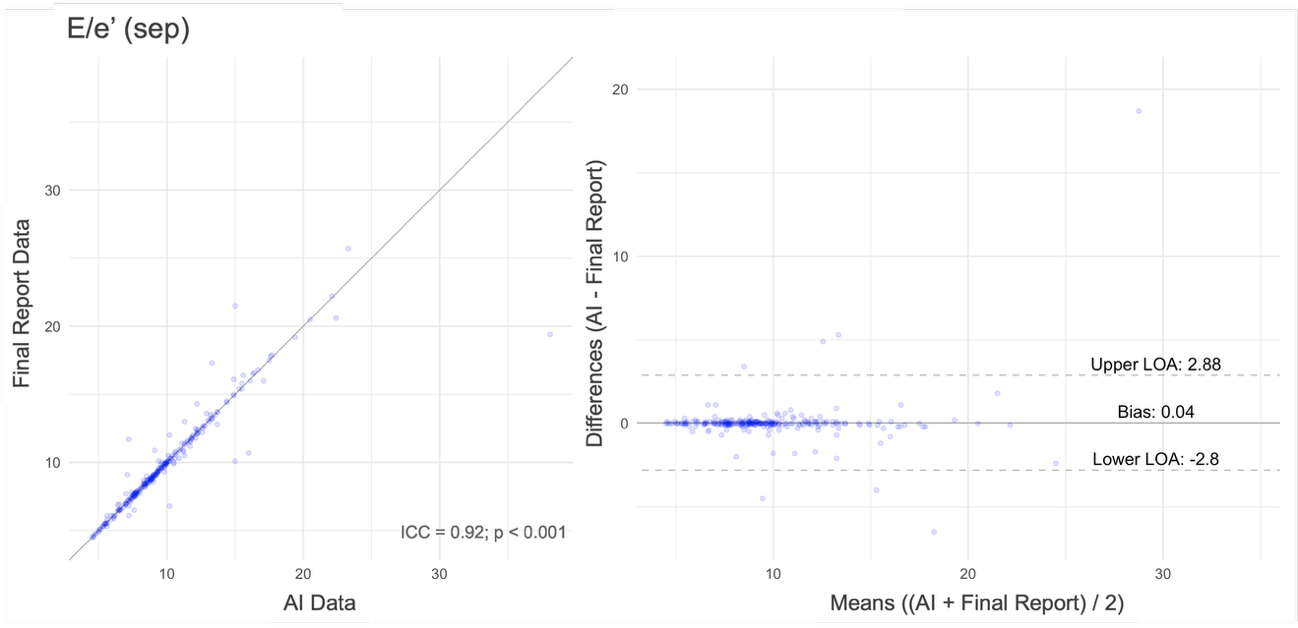

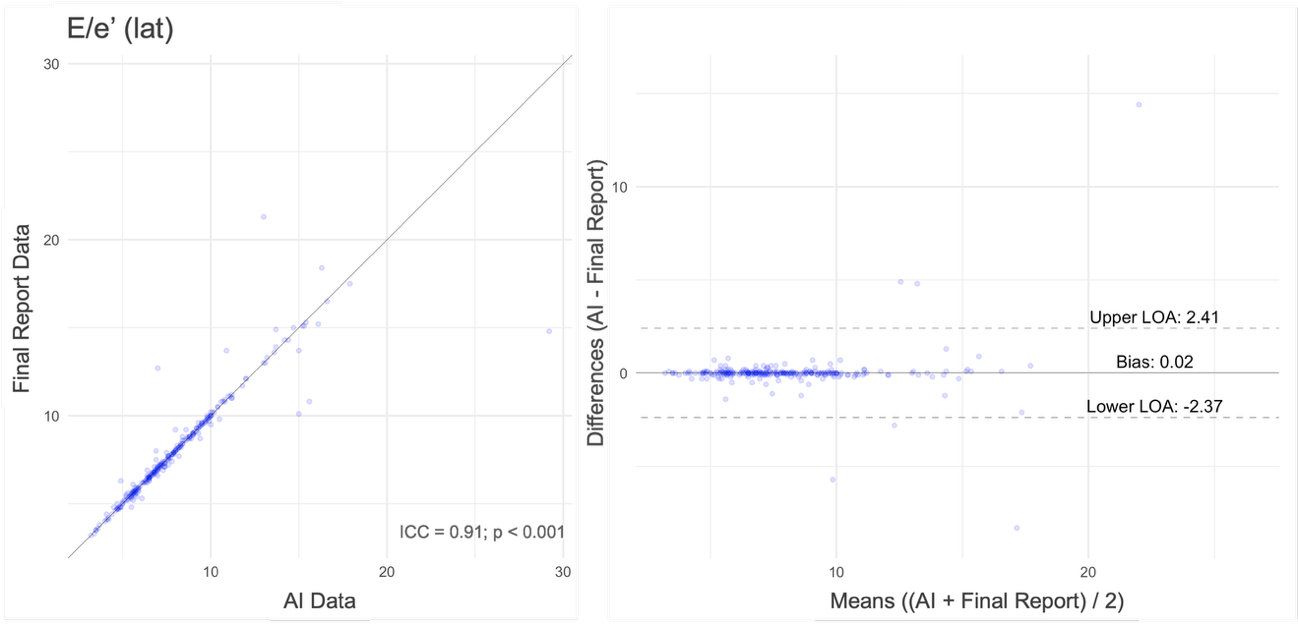

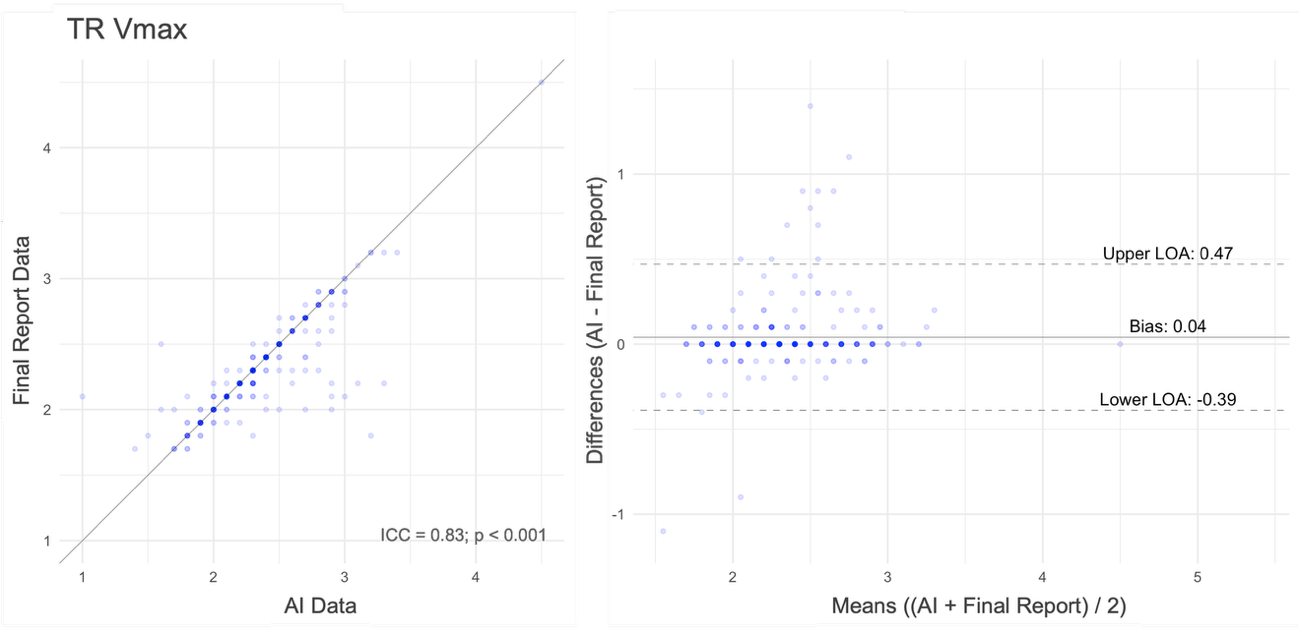

The scatter plots (left) demonstrate a strong correlation between AI and final report data. The Bland-Altman plots illustrate the mean differences (bias) and limits of agreement (LOA) between the two sets of measurements, with the bias being close to zero for each parameter.
